# Early Prediction of Parkinson’s Disease Progression by Integrating Research Cohort and Real-World Data Using Knowledge-Anchored Graph Learning

**DOI:** 10.64898/2026.07.07.26357483

**Authors:** Zuoyu Yan, Haoyang Li, Zhe Huang, Manqi Zhou, Wei-Ting Wang, Shujun Jiang, Mengying Zhang, Enrique Martinez-Nunez, Roberta Marongiu, Harini Sarva, Michael S. Okun, Jiayu Zhou, Chang Su, Fei Wang

## Abstract

Parkinson’s disease (PD) progression is highly heterogeneous. Deeply phenotyped longitudinal research cohorts have enabled characterization of PD progression trajectories. Early prediction of these progression patterns can help us better understand patient disease conditions and manage appropriately. However, the sample sizes of these cohorts are typically too small to build robust early predictors, and usually it is challenging to translate them to real-world patients because of the differences in the population as well as the information availability. In this paper we present MedStitcher, a graph-based machine learning framework that stitches individuals’ multimodal data across research cohorts and real-world data (RWD) using a biomedical knowledge graph-anchored architecture. This design enables predictive modeling under modality missingness and cross-dataset population heterogeneity. On the research cohort data combining PPMI and PDBP, MedStitcher achieved an AUROC of 0.819 ± 0.040 on predicting rapid PD progressors, outperforming existing machine learning approaches. Graph-based model interpretation revealed clinical and molecular drivers involving cognitive vulnerability, α-synuclein biology, vesicle trafficking and neuroinflammation. Importantly, MedStitcher-predicted rapid progressors in RWD cohort demonstrated elevated risks of dementia, falls, mild cognitive impairment, and gait impairment, which also enabled identification of early indicators of rapid PD progression in real world patient populations.

## Introduction

Parkinson’s disease (PD) is a progressive, neurodegenerative disorder underpinned by the loss of dopamine-producing neurons and degeneration of motor and non-motor basal ganglia circuitry. PD leads to disability and is associated with widespread central nervous system dysfunction^1,2^. PD is the second most prevalent neurodegenerative disease after Alzheimer’s disease and affects more than 11 million individuals worldwide, with projections exceeding 25 million by 2050^3^. Despite substantial developments in biomarker discovery and longitudinal cohort studies, no disease-modifying therapy has yet demonstrated consistent efficacy^4,5^. A critical challenge for therapeutic advancement is the profound inter-individual variability in disease progression, where patients demonstrate markedly different trajectories of motor deterioration, cognitive impairment, and other functional decline within the PD course. Without accurate early prediction of progression trajectories, clinical trial design remains inefficient, effect sizes have been diluted, and customized intervention has been challenging.

Over the past two decades, several multi-institutional longitudinal cohorts have been developed for PD, such as the Parkinson’s Progression Markers Initiative (PPMI)^6^ and the Parkinson’s Disease Biomarkers Program (PDBP)^7^, which include comprehensive patient information spanning motor scales, neuropsychological assessments, imaging biomarkers, and biospecimens. These cohorts have enabled research such as PD subtype discovery^8,9^ and prediction^10,11^. Among the many cohorts available internationally, specific enrollment criteria and information collected have varied. The sample sizes and data modalities available were also different. These limitations make it challenging to build robust machine learning models capable of producing generalizable insights.

In parallel, the widespread adoption of digital health infrastructures (such as the electronic health record system) has accumulated large-scale real-world data (RWD), inclusive of practice based information possibly useful for real world patients at scale^12^. Yet because RWD are collected primarily from clinical practice, they lack granular disease specific information such as PD-related clinical scales, harmonized biomarker measurements, and regular follow-up visits. From a machine-learning perspective, this makes it challenging to transfer the model built on research cohorts to RWD cohorts because of the discrepancies on the patient population and information availability.

In this paper, we propose to leverage biomedical knowledge graphs (BKGs) for bridging such gap. By encoding heterogeneous biomedical entities, such as clinical phenotypes, genes, medications, and pathways, within a relational network structure^13,14^, BKGs have the potential for semantically linking up datasets with different information modalities. In addition, recent advances in graph-based ML^15^ can further enable information propagation across the BKG to improve predictive modeling.

Specifically, we develop MedStitcher, a knowledge-enhanced graph machine learning framework for integrating heterogeneous biomedical data sets from both research and RWD cohorts to enable joint modeling. We applied MedStitcher on the problem of early predictive modeling of PD progression, where MedStitcher stitches together deeply phenotyped longitudinal PD research cohorts (PPMI and PDBP) with population-scale real-world data (All of Us^16^). Our technique adopted a BKG–anchored hybrid graph architecture which bridges and aligns patients across different cohorts with missing modalities with a shared progression manifold. By augmenting graph neural networks (GNNs) with graph test-time adaptation (GTTA)^17,18^, MedStitcher can handle cross-cohort population heterogeneity and support robust predictive modeling. Across five-fold nested cross-validation on pooled PPMI+PDBP data, MedStitcher achieved an AUROC 0.819 ± 0.040, outperforming gradient-boosted tree models and conventional GNN baselines. Model interpretation based on GNNExplainer^19^ revealed clinically and biologically coherent drivers of rapid progression in PD, spanning early motor and cognitive features as well as molecular pathways involving α-synuclein biology, vesicle trafficking and neuroinflammation. Importantly, MedStitcher detected probable rapid PD progressors in RWD using data collected within the first-year post PD diagnosis, which exhibited significantly elevated long-term risks of dementia, mild cognitive impairment, gait disorders, and recurrent falls during follow-up. Furthermore, examination of phenome-wide factors identified early indicators of rapid PD progressors, which were further validated using an independent RWD repository from the INSIGHT^20^ network. In this way, MedStitcher established a flexible framework for translating research-grade insights into real-world clinical settings, enabling population-level risk stratification and informing trial enrichment strategies.

## Results

Fig. 1 demonstrated the overall architecture of MedStitcher, which introduces a two-layer hybrid graph architecture in which a biomedical knowledge graph (BKG) bridges individuals across datasets with different information modalities (Methods, Fig. 1b). With this architecture, a graph neural network (GNN)^21^ learning procedure was conducted to jointly learn the patient representations through information aggregation from neighboring patients within the same dataset and patients in other datasets through paths in BKG that serve as bridges (Fig. 1c). To further address distribution discrepancies across heterogeneous data sets for improving model robustness, MedStitcher incorporates graph test-time adaptation (GTTA)^17^ to mitigate distribution shifts during inference. Overall, MedStitcher provides a new paradigm for integrating multimodal data across heterogeneous research and RWD cohorts to enable robust predictive modeling using graph machine learning.

**Figure 1.**
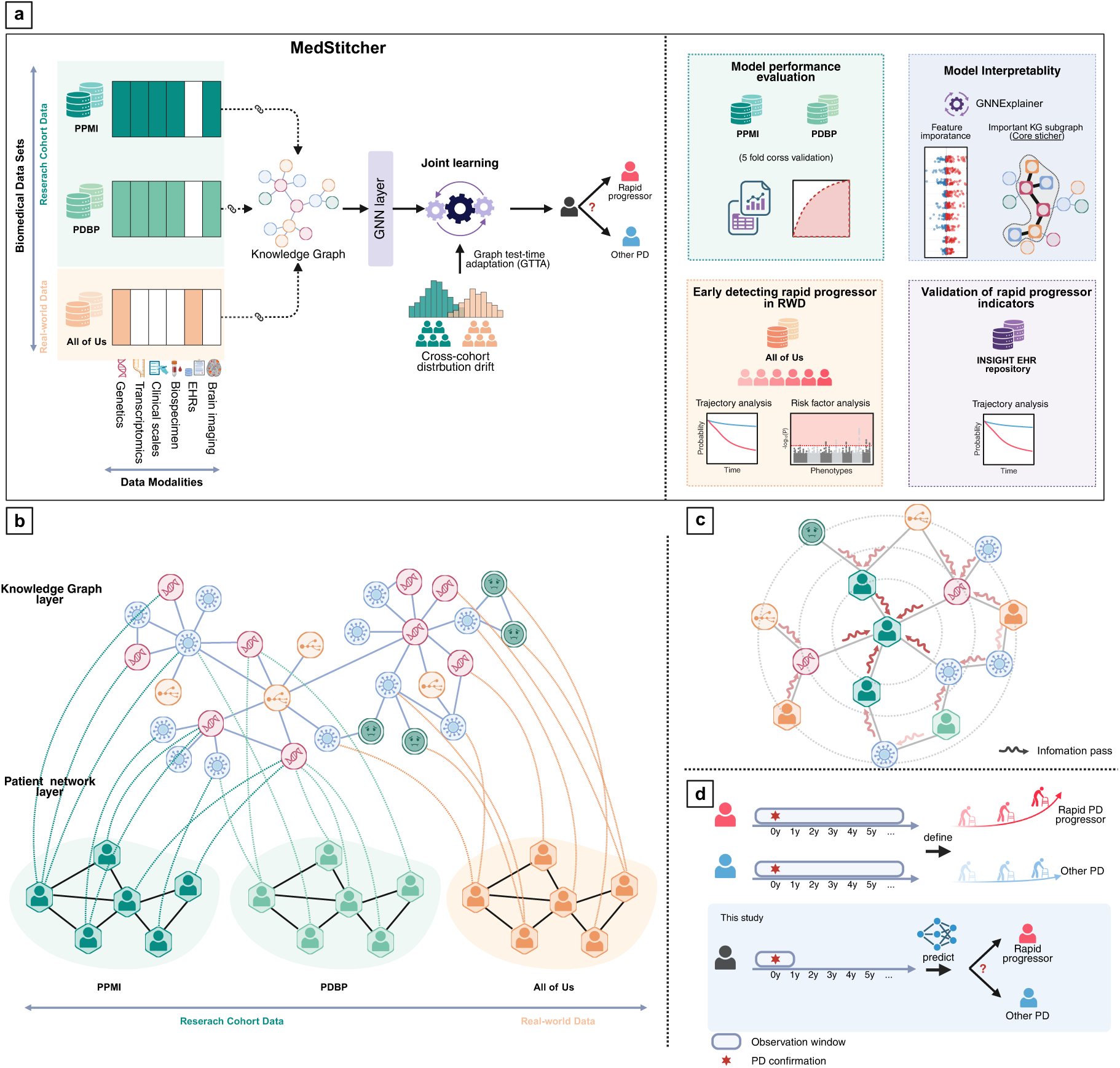
Illustration of MedStitcher for stitching research cohort data with real-world patient data (RWD) for early prediction of Parkinson’s disease progression. MedStitcher stitches multimodal patient data from research cohorts and RWD through a biomedical knowledge graph (BKG), generating a two-layer hybrid graph architecture (**a, b**). Within this architecture, a graph neural network (GNN) jointly learns patient representations by aggregating information directly from neighboring patients within the same dataset and indirectly from patients in other datasets through BKG nodes that serve as bridges (**c**). To mitigate cross-cohort feature misalignment and population shift, MedStitcher applies label-free graph test-time adaptation during inference, enabling robust transfer across datasets (**a**). Building on our prior identification of rapid PD progressors using more than 5 years of longitudinal PPMI and PDBP data, MedStitcher aims to predict PD progression subtype membership, distinguishing rapid progressors from other patients with PD (**d**). Model performance is evaluated in PPMI and PDBP, where ground-truth rapid progressor labels are available. GNNExplainer is used to assess feature importance and identify key bridge nodes. MedStitcher also predicted rapid PD progressors within the RWD and identify early indicators of rapid PD progression, which are subsequently evaluated in an independent RWD repository.

We particularly applied MedStitcher for addressing a challenging task for early prediction of PD progression subtypes. Two longitudinal, multi-center research studies of PD, the PPMI^6^ and PDBP^7^, that collect comprehensive clinical, biomarker, genetic and molecular data, and brain imaging of individuals, were leveraged in our study. We have also incorporated electronic health records and genetic data from the All of Us Research Program (AoU)^16^, a publicly available real-world dataset covering approximately half a million individuals in the United States. Our previous work has defined rapid progressors using longitudinal clinical assessments in PPMI and PDBP^9^. Here, our goal was to build a predictor that can distinguish rapid progressors from other PD patients early in the disease course (within one year after baseline) across both research cohorts (PPMI and PDBP), and transfer such predictor to the RWD cohort (All of Us), where the longitudinal clinical assessment information defining these subtypes may not be available (Fig. 2d, Extended Data Fig. 1). Detailed clinical characteristics of the patients from different cohorts are summarized in Table 1.

**Figure 2.**
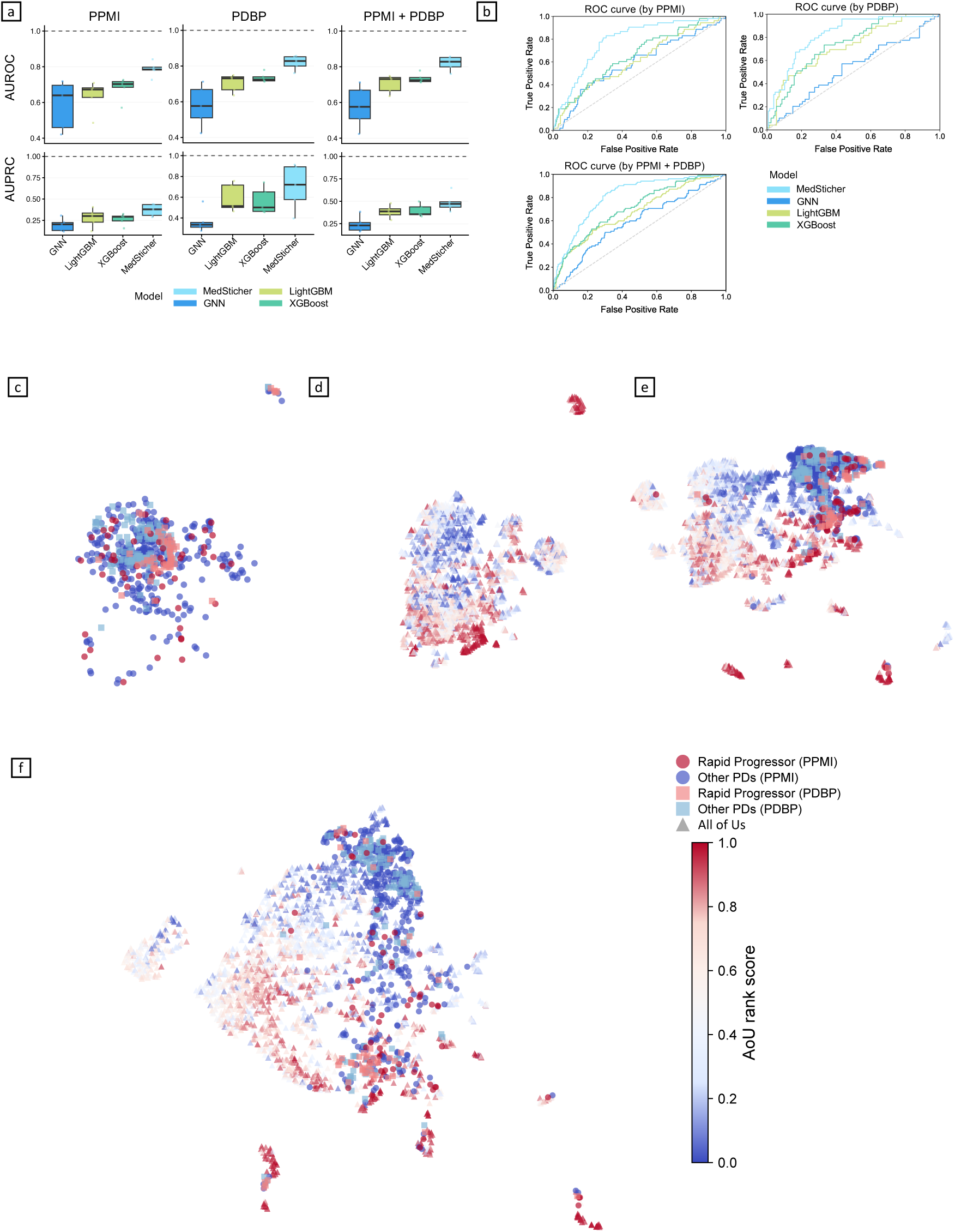
MedStitcher enables early prediction of Parkinson’s disease rapid progressors. **a, b.** Predictive performance of the models measured by receiver operating characteristic (ROC) curves as well as area under the ROC curve (AUROC) and area under the precision-recall curve (AUPRC). **c.** Uniform Manifold Approximation and Projection (UMAP) visualization of patient embeddings from the research cohorts PPMI and PDBP, derived from the original patient features. **d.** UMAP visualization of patient embeddings from the real-world dataset All of Us, derived from the original patient features. **e-f.** UMAP visualizations of embeddings for all patients with Parkinson’s disease from PPMI, PDBP and All of Us in the shared latent space learned by a conventional graph neural network (GNN) (**e**) and MedStitcher (**f**). Patients from PPMI and PDBP are colored as rapid progressor or other PDs. Patients from All of Us are colored according to the probability of belonging to the rapid progressors, as estimated by MedStitcher.

**Table 1.** Demographic characteristics of the studied Parkinson’s disease populations in PPMI, PDBP, and AoU.

| Characteristic | PPMI |  |  |  | PDBP<sup>a</sup> |  |  |  | All of Us<sup>b</sup> |  |  |  |
| --- | --- | --- | --- | --- | --- | --- | --- | --- | --- | --- | --- | --- |
|  | All PDs | Rapid Progressor | Other PDs | P value | All PDs | Rapid Progressor | Other PDs | P value | All PDs | Rapid Progressor (predicted) | Other PDs (predicted) | P value |
| # of patients | 405 | 53 | 352 | — | 176 | 49 | (N=127) | — | (N=2,561) | (N=513) | (N=2,048) | — |
| Age, mean (SD), yr | 61.5 (9.8) | 66.7 (8.9) | 60.8 (9.7) | <0.001 | 63.0 (10.0) | 65.2 (9.8) | 62.1 (10.0) | 0.079 | 71.2 (10.4) | 73.5 (9.1) | 70.6 (10.6) | <0.001 |
| Sex, N (%) |  |  |  |  |  |  |  |  |  |  |  |  |
| Female | 140 (34.6) | 10 (18.9) | 130 (36.9) | 0.010 | 73 (41.5) | 17 (34.7) | 56 (44.1) | 0.257 | 1,049 (41.0) | 210 (40.9) | 839 (41.0) | 0.720 |
| Male | 265 (65.4) | 43 (81.1) | 222 (63.1) |  | 103 (58.5) | 32 (65.3) | 71 (55.9) |  | 1,481 (57.8) | 295 (57.5) | 1,186 (57.9) |  |
| Race, N (%) | — | — | — | 0.718 | — | — | — | 0.405 | — | — | — | <0.001 |
| Black or African American | 6 (1.5) | 1 (1.9) | 5 (1.4) | — | 2 (1.1) | 0 (0.0) | 2 (1.6) | — | 151 (5.9) | 32 (6.2) | 119 (5.8) | — |
| Other race | 26 (6.4) | 5 (9.4) | 21 (6.0) | — | 7 (4.0) | 2 (4.1) | 5 (3.9) | — | 516 (20.1) | 153 (29.8) | 363 (17.7) | — |
| White | 373 (92.1) | 47 (88.7) | 326 (92.6) | — | 167 (94.9) | 47 (95.9) | 120 (94.5) | — | 1,894 (74.0) | 328 (63.9) | 1,566 (76.5) | — |
| Ethnicity, N (%) |  |  |  | 0.334 |  |  |  | 0.490 |  |  |  | <0.001 |
| Hispanic or Latino | 9 (2.2) | 2 (3.8) | 7 (2.0) |  | 6 (3.4) | 2 (4.1) | 4 (3.1) |  | 303 (11.8) | 100 (19.5) | 203 (9.9) |  |
| Not Hispanic or Latino | 396 (97.8) | 51 (96.2) | 345 (98.0) |  | 161 (91.5) | 43 (87.8) | 118 (92.9) |  | 2,154 (84.1) | 379 (73.9) | 1,775 (86.7) |  |
| Other ethnicity | 0 (0.0) | 0 (0.0) | 0 (0.0) |  | 9 (5.1) | 4 (8.2) | 5 (3.9) |  | 104 (4.1) | 34 (6.6) | 70 (3.4) |  |

<sup>a</sup>In PDBP, we included early PD (PD duration < 3 years at enrollment).
<sup>b</sup>In All of Us, the trained MedStitcher predicted a risk score for each PD patient belong to the rapid progressors. We ranked the patients based on the score and defined rapid progressors as the top 20% ranked individuals, while the remaining as other PDs.
Abbreviations: PDBP, Parkinson's Disease Biomarkers Program; PD, Parkinson's disease; PPMI, Parkinson's Progression Markers Initiative; SMD, standardized mean difference. PPMI and PDBP subtype labels (rapid progressor vs. other PDs) follow established clinical definitions<sup>9</sup>. All of Us subtype labels denote model-predicted phenotypes.

### MedStitcher improves early prediction of rapid PD progressors within PPMI and PDBP

We trained MedStitcher using a nested cross-validation strategy across the PPMI and PDBP cohorts. Model hyperparameters were selected within the inner five-fold cross-validation loop, and predictive performance was evaluated in the outer cross-validation loop. Because AoU data does not contain ground-truth “rapid PD progressor” information, AoU data were incorporated into the MedStitcher graph during both training and testing to support cross-dataset representation learning, but quantitative performance evaluation was restricted to PPMI and PDBP. We benchmarked MedStitcher against several widely used machine learning approaches, including LightGBM^22^, XGBoost^23^, and a graph neural network (GNN) approach^21^ (Methods).

As shown in Fig. 2a-b, across all evaluation settings, MedStitcher consistently outperformed competing methods, with an AUROC of 0.786 ± 0.041 and an AUPRC of 0.371 ± 0.070 in PPMI, AUROC of 0.841 ± 0.129 and an AUPRC of 0.599 ± 0.218 in PDBP, and AUROC of 0.819 ± 0.040 and an AUPRC of 0.486 ± 0.100 in pooled PPMI+PDBP hold-out sets. Notably, although AoU did not contain supervision information related to progression subtypes, incorporating AoU into the MedStitcher graph improved prediction performance, suggesting that unlabeled RWD can enhance cross-dataset representation learning (Supplementary Table 1).

We have also evaluated MedStitcher with the PPMI cohort for training and the PDBP cohort for testing. Due to the cohort differences and information discrepancy, conventional baselines transferred poorly, whereas BKG augmentation improved prediction and the full MedStitcher model achieved the best performance (AUROC = 0.759; AUPRC = 0.562; Supplementary Table 2). Ablation analyses showed that clinical features were the dominant contributor, with additional signal from genetics and more modest contributions from MRI and demographics (Extended Data Fig. 2, Supplementary Table 3). Sensitivity analyses showed that patient–patient similarity edges improved performance, while results remained stable across moderate graph sparsity and BKG message-passing-depth settings, supporting the robustness of the graph construction strategy (Extended Data Fig. 2, Supplementary Table 4).

To understand how MedStitcher improved prediction, we visualized patient embeddings using UMAP^24^ (Fig. 2c–f). In PPMI and PDBP, where ground-truth rapid PD progressor information was available, original patient features showed limited separation between rapid and non-rapid progressors (Fig. 2c-d). A similar pattern was observed with the GNN-only model without biomedical knowledge graph integration, where modest geometric separation improvement was accompanied by persistent cohort-specific clustering (Fig. 2e). Embeddings learned by the MedStitcher exhibited much better separation between rapid and non-rapid progressors in PPMI and PDBP (Fig. 2f). Embeddings of AoU individuals were also located in the same region as those embeddings derived from PPMI and PDBP, rather than forming an isolated cohort-specific cluster. These findings suggest that MedStitcher can learn a shared embedding manifold across heterogeneous datasets. Our findings suggested that, through message passing over the hybrid patient–BKG topology, MedStitcher aggregates disease-relevant signals that are not directly observed in patient features, improving separation of divergent progression trajectories across heterogeneous cohorts. This is also supported by theoretical results in Supplementary Note 1.

### MedStitcher reveals early clinical signatures and mechanistic drivers of rapid PD progression

To understand the biological and clinical interpretation with MedStitcher predictions, we applied GNNExplainer^19^ to the trained model. More specifically, GNNExplainer identified both patient-level feature attributions and compact BKG substructures that contributed to rapid PD progressor predictions.

At the patient layer, there are three major types of important features (Fig. 3a). First, cerebrospinal fluid biomarkers, including α-synuclein, Aβ42, and tau-related measures, were among the strongest predictors, consistent with evidence that mixed synuclein and Alzheimer-type pathology contributes to cognitive decline and accelerated progression in PD^25,26^. Second, baseline motor functional severity, including UPDRS II & III and Schwab and England ADL scores, as well as age, strongly influenced predictions, suggesting that reduced functional reserve is an early indicator of rapid progression. Third, subtle baseline cognitive deficits, including SDMT, semantic fluency, and HVLT performance, emerged as strong predictors, supporting that early cognitive vulnerability may precede overt progression in PD^27^.

**Figure 3.**
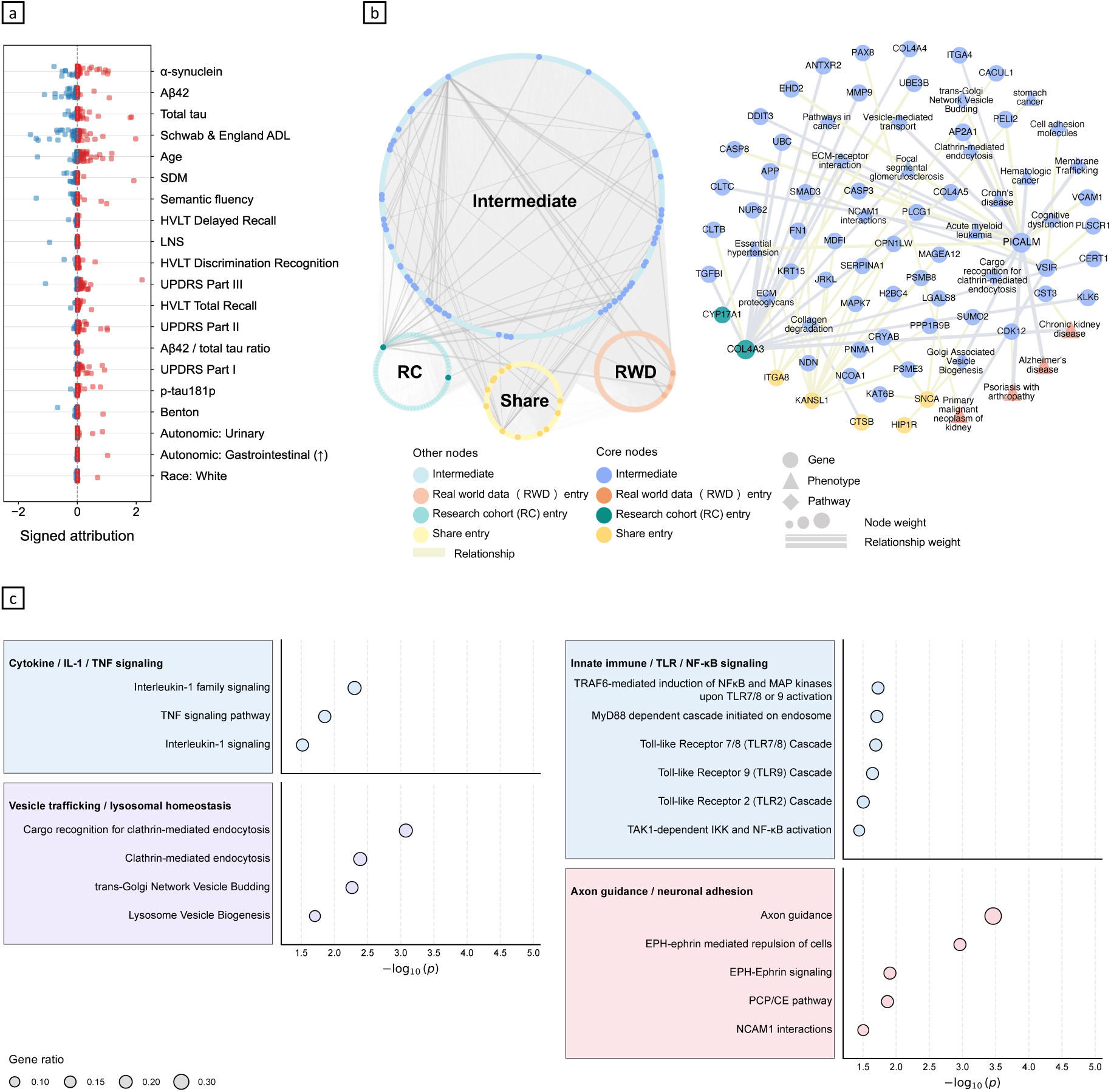
Model interpretation of MedStitcher predictions. Applying GNNExplainer to the trained MedStitcher, we assessed feature importance contributing to rapid PD progression prediction at the patient layer of the MedStitcher graph (**a**). At the BKG layer, GNNExplainer identified an explanatory subgraph functioning as a “core stitcher,” mediating knowledge and information transfer across heterogeneous datasets and supporting PD progression prediction (b). The left panel shows the core stitcher within the BKG, in which intermediate nodes bridge research cohort, real world data, and shared entry nodes; the right panel details the gene, phenotype and pathway entities within this core stitcher, with node and edge weights indicating explanatory importance. Pathway enrichment analysis of genes within the core stitcher subgraph further revealed biological pathways potentially underlying PD progression (**c**). Abbreviations: ADL, activities of daily living; KG, knowledge graph; PIGD, postural instability/gait difficulty; UPDRS, Unified Parkinson’s Disease Rating Scale.

At the BKG layer, we identified an important explanatory subgraph (Methods) that functioned as the “core stitcher”, mediating knowledge and information transfer across heterogeneous datasets and enabling improved PD progression prediction. This core stitcher subgraph consisted of 208 genes, 19 phenotypes, 25 biological pathways, and 207 relations within the BKG layer (Fig. 3b). Notably, genes represented the dominant bridging nodes, suggesting that molecular features serve as the primary anchors for cross-dataset information propagation.

A remarkable hub centered on *PICALM*, which was connected to *AP2A1*, clathrin-mediated endocytosis, vesicle trafficking pathways, cognitive dysfunction, and Alzheimer’s disease nodes, suggesting a model-attributed endocytic module that is biologically consistent with cognitive vulnerability in PD. More broadly, this cognitive-network interpretation is consistent with recent evidence conceptualizing PD as a somato-cognitive action network disorder^28^. Such observation biologically supported the emerging recognition of the role of impaired synaptic vesicle recycling and endolysosomal dysfunction in PD-related cognitive decline^29,30^. Another complementary module linked *SNCA* to endocytic regulators including *HIP1R*, *PSME3*, *KAT6B*, reinforcing the role of disrupted α-synuclein handling, proteostasis, and intracellular trafficking in PD progression^31–34^. We also identified a distinct module centered around *COL4A3*, which was connected to *FN1*, *COL4A5*, *MMP9*, chronic kidney disease, and extracellular matrix pathway nodes^35,36^. Rather than representing a canonical PD-specific mechanism, this network reflected broader systemic vulnerability, extracellular matrix remodeling, and multimorbidity burden that could accelerate long-term functional decline. Pathway enrichment analysis based on genes of the core subgraph further identified four recurrent biological programs including inflammatory cytokine signaling, innate immune/TLR/NF-κB pathways, vesicle trafficking and lysosomal homeostasis, and axonal guidance and neuronal adhesion pathways (Fig. 3c).

### MedStitcher identifies probable rapid progressor in real-world data (RWD)

Unlike PPMI and PDBP, the AoU dataset lacks PD-specific longitudinal assessments required to define progression subtypes including the rapid progressors^9^. However, after training, the MedStitcher generated an individual-level risk score representing each patient’s probability of belonging to the rapid progression subtype within this cohort. First, in consistence with the rapid progressors originally defined in PPMI and PDBP, the predicted rapid progressors were significantly older than other PD patients at disease onset (age 73.5 ± 9.1 vs 70.6 ± 10.6 years; p-value < 0.001). (Table 1) We next evaluated whether the predicted risk can be translated into accelerated real-world disease progression by examining time to advanced PD outcomes. To this end, we assessed motor outcomes, including gait disorders and falls, and cognitive outcomes, including dementia and mild cognitive impairment. Kaplan–Meier analyses models were used to estimate time to first event, while recurrent falls were evaluated using a recurrent-event Cox model, adjusted for covariates including age, sex, race and ethnicity. As shown in Fig. 4a-d, the predicted rapid progressors exhibited consistently worse longitudinal outcomes across all endpoints, including a 30% higher risk of developing dementia (P = 0.015), a 24% higher risk of mild cognitive impairment (P = 0.049), a 53% higher risk of gait disorder (P = 0.014), and a significant higher risk of recurrent falls (adjusted HR = 2.97, 95% CI [1.25, 7.05], P = 0.014), compared with predicted non-rapid individuals. Notably, the explanatory KG derived after transfer to RWD was distinct from the primary core stitcher, forming a shared intermediate core organized around immune/HLA signaling, lysosomal–proteostatic biology and neuropsychiatric burden (Extended Data Fig. 3a,b).

**Figure 4.**
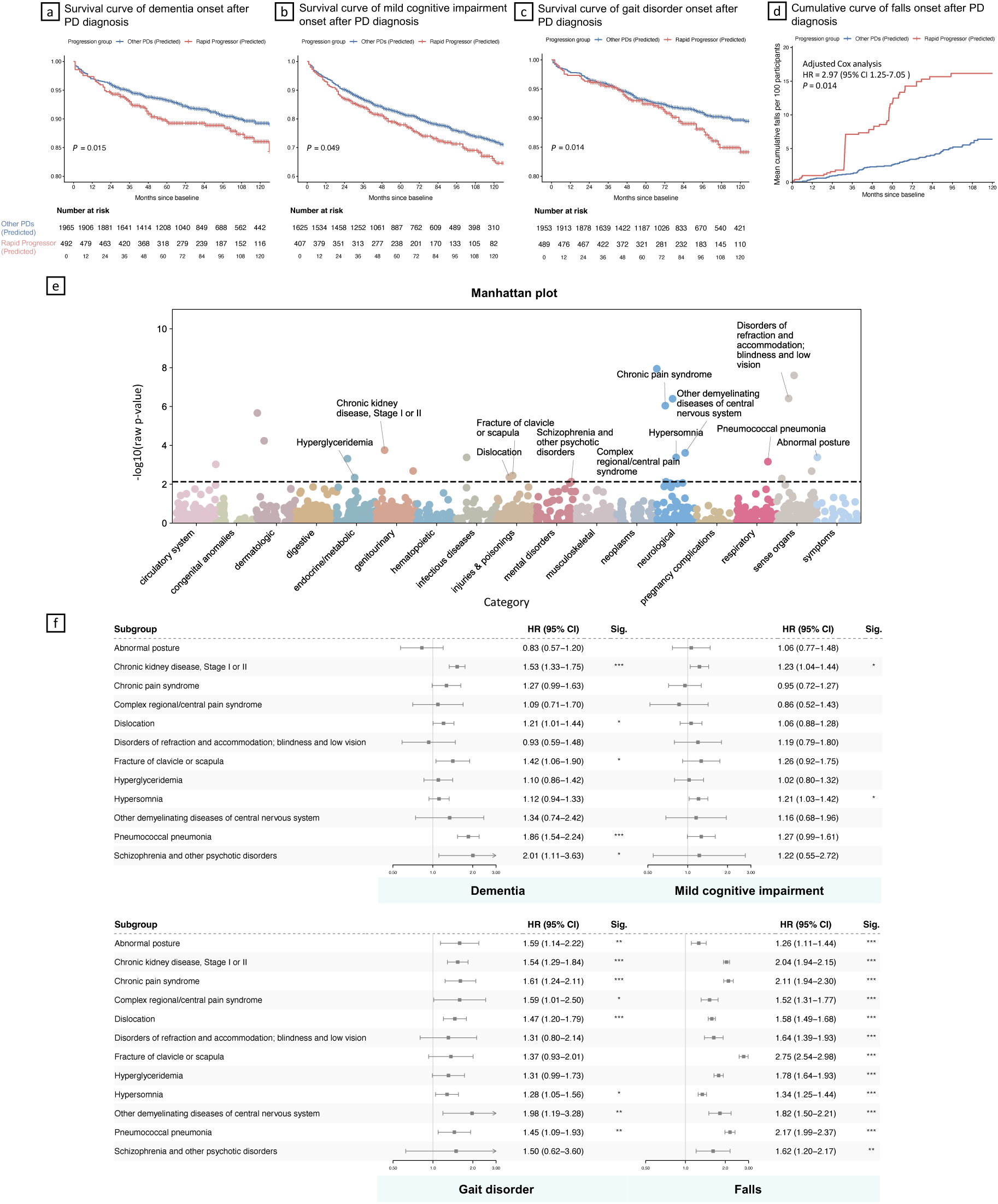
Identification of Parkinson’s disease progression indicators. a–d, Longitudinal clinical validation of MedStitcher-predicted rapid progressors in the All of Us PD cohort. **a–c**, Kaplan–Meier event-free survival curves for incident dementia (a), mild cognitive impairment (b) and gait disorder (c), comparing MedStitcher-predicted rapid progressors with other PD patients. **d**, Mean cumulative function curve for recurrent falls, showing cumulative fall burden per 100 participants after PD diagnosis, the group effect was estimated using an Andersen–Gill recurrent-event Cox model adjusted for age, sex, race and ethnicity, with robust standard errors clustered by participant. Predicted rapid progressors were defined as the top 20% of individuals ranked by MedStitcher-predicted rapid-progression risk. **e.** Manhattan plot visualizing phenome-wide potential indicators of rapid PD progression. Phenotypes were collected within the baseline window (before one year post PD diagnosis). Phecodes were derived from ICD codes using established ICD-to-phecode mappings and tested by covariate-adjusted logistic regression (age, sex, race and ethnicity), including all phecodes^62^ observed at least once (1732 phecodes); points are grouped by phecode category and the dashed line denotes the Bonferroni significance threshold (P < 0.05 / 1732). **f.** External validation in the INSIGHT real-world data repository. For each indicator, we split the PD patients in INSIGHT into carriers and non-carriers based on baseline records, and fitted Cox proportional hazards models, adjusting for covariates, to test its associations with 10-year follow-up outcomes. Hazard ratios were visualized using forest plot with 95% confidence intervals.

### MedStitcher-transferred PD progression signals identify early indicators of rapid progression

We next sought to identify early clinical indicators of rapid progression using routine clinical records in AoU. We examined phenotypes recorded from PD diagnosis through one year after diagnosis and identified baseline phenotypes significantly enriched among predicted rapid progressors. This resulted in a list of potential early indicators of rapid PD progression (Fig. 4e).

To validate these indicators, we used INSIGHT clinical research network, an independent large-scale RWD repository comprising 40,158 eligible patients with PD, for external validation. For each phenotype, the PD patients were divided into exposed and non-exposed groups, where exposure was defined by whether the condition was present within the same baseline window from PD diagnosis to 1 year after diagnosis. Then Cox models were used to test PD progression rates measured by onset of above-mentioned outcomes including dementia, mild cognitive impairment, gait disorder, and falls, during the 10-year follow-up after PD confirmation. The models adjusted for covariates including age, sex, race and ethnicity.

Remarkably, baseline onset of chronic kidney disease (CKD) among the PD patients emerged as the most consistent signal, remaining significantly associated with all four progression outcomes: dementia (HR 1.53, 95% CI 1.33–1.75), mild cognitive impairment (HR 1.23, 95% CI 1.04–1.44), gait disorder (HR 1.54, 95% CI 1.29–1.84) and recurrent falls (HR 2.04, 95% CI 1.94–2.15) (Fig. 4f). Other phenotypes including dislocation, clavicle/scapular fracture, hypersomnia, pneumococcal pneumonia, and psychotic disorders, were also associated with both motor and cognitive outcomes, suggesting a broader vulnerability state spanning injury susceptibility, sleep dysfunction, respiratory frailty, and neuropsychiatric burden (Fig. 4f). In addition, abnormal posture, chronic pain syndromes, visual disorders, hyperglyceridemia, and demyelinating disorders were primarily associated with gait-related outcomes, defining a more mobility-centered risk axis (Fig. 4f).

## Discussion

Complex diseases such as PD arise from intertwined molecular, biological, and clinical processes unfolding across multiple scales^37^. Although the rapid growth of multimodal biomedical data—from deeply phenotyped research cohort studies to population-scale real-world patient data (RWD)—has created unprecedented opportunities for disease modeling, these datasets remain highly fragmented across institutions, generated under heterogeneous protocols, and characterized by substantial differences in modality availability and granularity. Such fragmentation limits the development of robust and generalizable AI and machine learning models, as most prior approaches rely on single datasets or require highly harmonized data structures. Here, we present MedStitcher, a knowledge-enhanced machine learning framework designed to bridge this divide by integrating deeply phenotyped research cohorts with large-scale RWD for early prediction of PD progression.

Building upon our prior identification of PD progression subtypes using longitudinal clinical assessments within full-PD course in research cohort studies^9^, MedStitcher addresses two major translational challenges: predicting progression trajectories, particularly the rapid progressors, early in the disease course and extending research-derived progression insights to real-world populations that lack granular PD-specific phenotyping. By stitching together research cohort studies (PPMI, PDBP) and RWD (All of Us) through a hybrid graph architecture based on BKG, MedStitcher enables robust prediction of rapid progressors despite substantial modality missingness and distributional heterogeneity across datasets.

A major conceptual contribution of this work is demonstrating that, within the MedStitcher framework, a BKG can function as an effective bridge between fragmented biomedical ecosystems to improve machine learning modeling. Rather than treating missing modalities as a limitation that requires imputation or exclusion, MedStitcher leverages structured biological relationships among genes, phenotypes, pathways, and diseases within the BKG architecture to propagate disease-relevant information across datasets. This enables building graph-based machine learning especially graph neural networks to embed patients with highly heterogeneous feature availability into a shared progression manifold. More broadly, our results highlight the potential of MedStitcher to reconcile heterogeneous biomedical datasets and translate research-grade insights into scalable tools for disease and/or progression modeling in complex neurodegenerative disorders.

Our approach has important clinical and translational implications for PD research and therapeutic development. Early identification of patients at risk for rapid progression remains a critical unmet need in PD, as clinical trials for disease-modifying therapies are frequently hindered by marked heterogeneity in disease trajectories, which can dilute treatment effects and obscure therapeutic signals. By enabling scalable risk stratification across both deeply phenotyped research cohorts and grounding into population-scale real-world clinical data, MedStitcher provides a practical strategy for identifying high-risk individuals early in the disease course. Such capability may facilitate more efficient trial enrichment by prioritizing patients more likely to exhibit measurable progression within the trial window, thereby improving statistical power and reducing sample size requirements. Beyond trial design, early identification of fast progressors may also inform clinical management by supporting more proactive monitoring and earlier implementation of supportive interventions aimed at mitigating downstream complications such as cognitive decline, falls, and functional impairment. Importantly, because the framework enables translation of research-cohort–derived progression trajectories into real-world clinical populations, it offers a path toward integrating predictive progression modeling into routine clinical settings.

Beyond predictive performance, MedStitcher generated biologically meaningful insights into mechanisms underlying rapid PD progression. Model interpretability analyses converged on early cognitive vulnerability, baseline motor-functional burden, and mixed neurodegenerative biomarker signatures involving α-synuclein, amyloid, and tau biology. At the molecular level, the identified “core stitcher” network highlighted biologically coherent pathways involving vesicle trafficking, endolysosomal dysfunction, α-synuclein handling, neuroinflammation, and neuronal adhesion. In particular, PICALM-centered endocytic pathways and SNCA-linked trafficking networks reinforce emerging evidence that disruption of proteostasis and intracellular trafficking may contribute to accelerated progression, particularly cognitive decline. These findings suggest that rapid progression in PD may emerge from interacting molecular programs rather than a single dominant biological pathway.

MedStitcher enabled discovery of early progression indicators in real-world clinical settings. Based on the predicted rapid progressors, phenome-wide analyses in All of Us identified chronic kidney disease, hypersomnia, fractures, psychotic disorders, and respiratory conditions as potential early indicators of rapid progression in PD. Importantly, these signals were validated over an independent large-scale RWD repository, INSIGHT, where they remained associated with long-term outcomes relevant to PD progression. These observations suggest that early manifestations of rapid progression may extend beyond traditional neurological symptoms and reflect broader multisystem vulnerability involving metabolic dysfunction, frailty, sleep disturbances, neuropsychiatric burden, and injury susceptibility. While these associations require further mechanistic investigation, they may offer clinically actionable signals for earlier monitoring and intervention.

Several limitations should be acknowledged. First, progression subtype labels were derived from prior unsupervised data-driven analyses rather than prospective clinical adjudication. Second, outcome definitions in RWD relied on diagnosis codes and clinical proxies that may introduce measurement error. Third, although we validated findings across multiple independent datasets, the RWD were derived from U.S.-based healthcare systems, which may limit generalizability to other populations. Finally, prospective validation will be necessary before clinical deployment.

## Methods

### Study populations

**Parkinson’s Progression Markers Initiative (PPMI),** launched in 2010 and sponsored by the Michael J. Fox Foundation, is an international and multi-center observational study dedicated to identifying biomarkers indicative of PD progression^6^. The present study included the following participants in the PPMI cohorts: *de novo* PD participants (diagnosed with PD within the last 2 years and untreated at enrollment), HCs, and individuals with SWEDD (scans without evidence of dopaminergic deficit). More information about PPMI participants has been described elsewhere^6^. The PPMI study protocol was approved by the institutional review board of the University of Rochester (NY, USA), as well as from each PPMI participating site. Data used in the preparation of this article were obtained from the PPMI database (www.ppmi-info.org/access-data-specimens/download-data), RRID:SCR_006431. From PPMI, we included individuals diagnosed with PD at baseline for analysis.

**Parkinson Disease Biomarkers Program (PDBP),** established in 2012 and funded by the National Institute of Neurological Disorders and Stroke (NINDS), is an observational study for advancing comprehensive PD biomarker research^7^. The present study included the participants with PD and HCs in the PDBP cohort. More information about the PDBP participants has been described elsewhere^7^. The study protocol for each PDBP site was approved by institutional review board of each participating site. Data of the PDBP cohort were obtained via the Accelerating Medicines Partnership Parkinson’s disease (AMP-PD) Knowledge Platform (http://amp-pd.org) under AMP-PD Data Use Agreement and PDBP webpage (https://pdbp.ninds.nih.gov). From PDBP, we included early PD individuals (symptom duration at enrollment < 3 years).

**All-of-Us**^16^, a nationwide initiative in the United States designed to enroll and follow more than one million participants to advance precision medicine and population health research. At the time of the data release used in this study, the Researcher Workbench contained data for approximately half a million participants. We analyzed the data using the AoU Controlled Tier CDR v8 dataset, which allows authorized users to conduct research projects without requiring individual Institutional Review Board (IRB) review. We identified participants with PD using diagnosis codes including G20, 332.0, and 49049000. Patients whose age < 50 years at the first PD diagnosis event were excluded for analysis.

**INSIGHT Clinical Research Network (CRN)**^20^, supported by the Patient-Centered Outcomes Research Institute (PCORI). In the INSIGHT extract used for this study, data were drawn from five New York City top academic medical centers and covered longitudinal records for over 15 million patients. These centers include Weill Cornell Medicine (WCM), Albert Einstein School of Medicine/Montefiore Medical Center, Columbia University Irving Medical Center, Icahn School of Medicine/Mount Sinai Health System, and NYU Grossman School of Medicine/NYU Langone Health. We used de-identified, longitudinal EHRs of patients in INSIGHT CRN between general 2010 to September 2024. Use of INSIGHT CRN data was under the approval of IRB at WCM (IRB number 24-07027736). We identified participants with PD using diagnosis codes including G20 and 332.0. Patients whose age < 50 years at the first PD diagnosis event were excluded for analysis. After applying these criteria, the final INSIGHT validation cohort included 40,158 patients with PD.

### Study design

Using over 5 years longitudinal clinical assessments from the PPMI and PDBP data with unsupervised deep learning, we have previously defined and validated three PD subtypes, each showing a unique progression trajectory within the disease course, including inching, moderate, and rapid progression subtypes^9^. In this study, we formulated the goal of PD progression prediction as **1)** distinguishing the rapid progressors (rapid progression subtype) from the other PD population within a short period (1 year window) after enrollment in PPMI and PDBP; **2)** and translating the PD progression subtypes into real-world settings by identifying the possible rapid progressors within the real-world data, i.e., All-of-Us.

### Data modality and data preparation

#### Clinical scales & CSF biomarkers from PPMI & PDBP

We used participant’s longitudinal data in diverse clinical assessments. Specifically, we used motor manifestation data including Movement Disorders Society–revised Unified Parkinson’s Disease Rating Scale (MDS-UPDRS) Parts II and III^38^ and Schwab-England activities of daily living score, as well as non-motor manifestation data including MDS-UPDRS Part I, Scales for Outcomes in Parkinson’s disease-Autonomic (SCOPA-AUT)^39^, Geriatric depression scale (GDS)^40^, Questionnaire for Impulsive-Compulsive Disorders in Parkinson’s disease (QUIP)^41^, State-Trait Anxiety Inventory (STAI)^42^, Benton Judgment of Line Orientation (JOLO)^43^, Hopkins Verbal Learning Test (HVLT)^44^, Letter-number sequencing (LNS), Montreal Cognitive Assessment (MoCA)^45^, Semantic verbal-language fluency test^46^, Symbol–Digit Matching (SDM)^47^, Epworth Sleepiness Score (ESS)^48^, REM sleep behaviour disorder (RBD)^49^, and Cranial Nerve Examination^50,51^.

We also used CSF biospecimen data including *α* -synuclein measured by enzyme-linked immunosorbent assay^52^, amyloid-beta1–42 (*Aβ*-42), phosphorylated Tau protein at threonine 181 (P-tau), and total tau protein (T-tau) measured by INNO-BIA AlzBio3 immunoassay. Following the previous studies^53,54^, we also evaluated *Aβ* -42/P-tau, *Aβ* -42/T-tau, *Aβ* -42/ *α* -synuclein, P-tau/ *α* - synuclein, T-tau/*α*-synuclein, and P-tau/T-tau levels. Availability of above clinical assessment and CSF biomarker variables can be found in in the Supplementary **Table 5**.

#### Neuroimaging from PPMI & PDBP

We obtained high resolution T1-weighted 3 T MRI data of participants collected at baseline and 1-year follow-up in the PPMI and PDBP. For each individual, we calculated 1-year brain atrophy measured by average cortical volume in 34 brain regions of interests (ROIs), defined by the Desikan-Killiany atlas (averaged over the left and right hemispheres)^55^. For each ROI, 1-year cortical atrophy was defined as the percentage change in cortical volume from baseline to 1-year follow-up, calculated as (1-year follow-up volume − baseline volume) / baseline volume × 100. ROI volumes were averaged across the left and right hemispheres according to the Desikan–Killiany atlas.

#### Whole genome sequencing (WGS) data from PPMI, PDBP & All-of-Us

We used WGS data of individuals.

#### Whole blood RNA-seq from PPMI & PDBP

We used whole-blood bulk RNA-seq data of participants.

#### Electronic health records (EHRs) from All-of-Us & INSIGHT^16,20^

We used diagnosis records both All-of-Us and INSIGHT EHRs. Data were collected within an observation window defined as from 5-year before to 1-year after the first PD diagnosis date. We excluded diagnosis codes with prevalence <0.5% to reduce sparsity and noise.

### MedStitcher: Stitching diverse datasets with a two-layer hybrid graph architecture

MedStitcher adopts a two-layer hybrid graph architecture including a patient layer formulated and a knowledge graph layer.

#### Patient layer

Patient–patient edges were constructed using a K-nearest-neighbor graph (K = 40) based on Euclidean-distance similarity; when available, MRI-derived representations were used to refine edge weights without imputing missing MRI measurements. In particular, we use the features that cannot be connected to the KG to compute the similarity between patient nodes. These features include the demographic feature, Movement Disorders Society–revised Unified Parkinson’s Disease Rating Scale (MDS-UPDRS) Parts II and III^38^, Schwab-England activities of daily living score, MDS-UPDRS Part I, Scales for Outcomes in Parkinson’s disease-Autonomic (SCOPA-AUT)^39^, Questionnaire for Impulsive-Compulsive Disorders in Parkinson’s disease (QUIP)^41^, Benton Judgment of Line Orientation (JOLO)^43^, Hopkins Verbal Learning Test (HVLT)^44^, Letter-number sequencing (LNS), Montreal Cognitive Assessment (MoCA)^45^, Semantic verbal-language fluency test^46^, Symbol–Digit Matching (SDM)^47^, Cranial Nerve Examination^50,51^, *α* -synuclein measured by enzyme-linked immunosorbent assay^52^, amyloid-beta1–42 (*Aβ*-42), phosphorylated Tau protein at threonine 181 (P-tau), and total tau protein (T-tau) measured by INNO-BIA AlzBio3 immunoassay, *β*-42/P-tau, *Aβ*-42/T-tau, *Aβ* -42/*α* -synuclein, P-tau/*α* -synuclein, T-tau/*α* -synuclein, and P-tau/T-tau levels. When available, the MRI-derived representations were used to refine edge weights.

#### Knowledge graph layer

We used a comprehensive BKG we built previously, termed the integrative biomedical knowledge hub (iBKH)^56^. iBKH has integrated and harmonized data from 18 publicly available biomedical knowledge sources, including biomedical ontologies, manually curated knowledge bases, existing biological knowledge graphs, and other biomedical data. It includes approximate 2.4 million biomedical entities (nodes) of 11 types, such as genes, diseases, symptoms, pathways, drugs, etc.) as well as over 48 million relations of 45 relation types, such as disease-associated-with_gene, disease-associated-with_disease, gene-bind-gene, etc.).

Give the huge scale of iBKH, incorporating the entire iBKH could introduce substantial noise and unnecessary complexity for PD-specific modeling. Therefore, we constructed a PD-focused subgraph by extracting entities and relations within the two-hop neighborhood of the “Parkinson’s disease” entity. Specifically, we retained entities of four types, including genes, diseases, symptoms, and pathways, along relations among them. To further improve data quality, low-confidence relations (relations extracted solely using machine learning methods in iBKH construction procedure) were excluded. The resulting PD-centric subgraph comprised 6,911 gene nodes, 588 disease nodes, 640 pathway nodes, and 248 symptom nodes, forming the biomedical knowledge backbone for subsequent graph-based learning.

#### Connecting patient nodes with entities within the knowledge graph layer

Our goal is to establish connections between patient nodes and entity nodes within the knowledge graph layer so that patients from heterogeneous datasets—even when data modalities are partially missing or unaligned—can be represented within a unified graph architecture. This design enables information propagation across datasets using graph-based learning algorithms.

1) Linking patient nodes to gene nodes using genetic data (genetic patient-gene relation construction). To incorporate genetic variation while mitigating the high dimensionality and small effect sizes of individual single-nucleotide polymorphisms (SNPs), we restricted the analysis to loci previously reported in genome-wide association studies (GWAS) related to Parkinson’s disease (PD). Specifically, we curated loci associated with PD risk, PD progression, PD-related pathology (e.g., α-synuclein measurements), and other neurodegenerative disorders—including Alzheimer’s disease, Huntington’s disease, dementia, and Lewy body dementia—from the GWAS catalog^57^. GWAS studies focusing on secondary or context-specific traits—such as young adult-onset parkinsonism, age at onset of PD, pesticide exposure measurement, age at diagnosis, Lewy body measurement, motor function measurement, drug-induced dyskinesia or response to levodopa, impulse control disorder, and mortality—were excluded. After filtering, we obtained 187 loci (referred to as PD risk loci). These loci were mapped to genes according to the reported HGNC symbols in the GWAS catalog and defined as gene entry nodes. A genetic patient-gene relation was constructed between a patient and a gene if the patient carried a risk variant within a locus mapped to that gene.

2) Linking patient nodes to gene nodes using gene expression data (transcriptomic patient-gene relation construction). RNA-seq data were used to connect patient nodes to gene entry nodes. Gene annotations were obtained from UniProt^58^, and only protein-coding genes were included. To reduce dimensionality and prioritize disease-relevant genes, differential expression analysis between PDs and normal controls was performed using DESeq2^59^. Genes meeting the criteria of |log₂ fold change| ≥ 0.2 and false discovery rate (FDR) < 0.05 were retained and added to the gene entry node list. A transcriptomic patient-gene relation was constructed between each patient and each of these gene nodes, with edge weight representing normalized gene expression level.

3) Linking patient nodes to disease/symptom nodes using clinical assessments (patient-phenotype relation construction). We curated a set of phenotypes based on the PD clinical assessments. These include: Hypersomnia, Depressive disorder, Anxiety state, Generalized anxiety disorder, REM sleep behavior disorder, Chronic pain syndrome, and Hallucinations. Each phenotype was represented as a phenotype entry node, and edges were established between patient nodes and phenotype nodes according to the corresponding assessment measurements.

4) Linking patient nodes to disease/symptom nodes using EHRs (patient-phenotype relation construction). We incorporated diagnosis information from EHRs in the All-of-Us dataset. In All-of-Us, diagnoses are documented using both ICD and SNOMED codes. To address this, SNOMED codes were first mapped to ICD-10-CM codes using the SNOMED CT to ICD-10-CM Map obtained from the Unified Medical Language System (UMLS)^60^. Subsequently, ICD-9 and ICD-10 codes were mapped to disease or symptom entities (i.e., phenotypes) using the dictionary provided by iBKH. These entities were also defined as phenotype entry nodes. A patient node was connected to a phenotype entry node if the corresponding diagnosis code was present in the patient’s EHR record.

### MedStitcher: deep graph neural network architecture

Our model used a multi-relational graph neural network with type-specific feature encoders followed by edge-aware graph propagation. Specifically, nodes from different data sources were first projected into a shared latent space using separate multilayer perceptron encoders, such that each node type was transformed into a common embedding of dimension *d* before graph aggregation. To be specific, let **x***_i_* denote the raw feature vector of node *i*, and let *m_i_* ∈ {0,1,2} indicate its node type (research cohort patient nodes, real-world data patient nodes, and knowledge graph nodes). The initial hidden representation was defined as 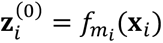, where *f_m_* denotes the corresponding type-specific encoder. This design allowed the model to preserve source-specific feature structure while aligning heterogeneous patient and knowledge-layer entities into a unified representation space. Graph propagation was then performed using a stack of graph attention layers by default, with graph convolution available as an alternative backbone. For each edge (*i*, *j*), the model used edge attributes as type-aware relational features, enabling attention weights and message passing to depend not only on neighboring node states but also on the semantic identity and strength of the connection. In the graph attention setting, node representations were iteratively updated as 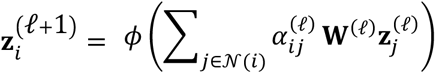 where 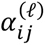 is an edge-aware attention coefficient conditioned on the pairwise interaction and the edge feature vector^21^. In the graph attention backbone, the attention coefficient *α_i_*_)_ was computed internally by an edge-aware graph attention operator as a normalized function of the transformed source-node embedding, target-node embedding and edge attribute. Thus, message passing was conditioned jointly on neighboring node states and relation-specific edge semantics, rather than on node features alone. **W**^(ℓ)^ is a learnable projection, and *φ*(⋅) denotes nonlinear activation followed by optional batch normalization, residual connection and dropout. After the final message-passing layer, node-level logits were obtained through a linear classifier applied to the learned hidden representation. In this framework, prediction for each patient aggregated information from three complementary sources: the patient’s own clinical features through the type-specific encoder, neighboring patients connected through the similarity layer, and biologically or clinically linked entities connected through the knowledge graph. The resulting architecture therefore supported relation-aware information fusion across heterogeneous modalities while retaining explicit edge semantics during propagation.

### MedStitcher: Graph test-time adaptation to address population shift

To mitigate data shift issue across multiple datasets, we applied a label-free graph test-time adaptation (GTTA) procedure, a label-free machine learning framework^17,18^, at inference to align heterogeneous datasets and enable integration across differing distributions. For each mini-batch *B* of target (seed) patients (maximum batch size = 2,048), we constructed a test subgraph by layer-wise neighbor sampling with a three-hop fanout of [50, 30, 20]. We then performed a lightweight episodic adaptation by updating only the batch-normalization (BN) affine parameters Θ_BN_ = {*γ_ℓ_*, *β_ℓ_*}*_ℓ_*, while keeping all remaining model weights fixed. Adaptation was driven by entropy minimization on the predicted probabilities *p_i_* = *σ*(*z_i_*) for seed nodes *i* ∈ *B*, using the mean binary entropy objective

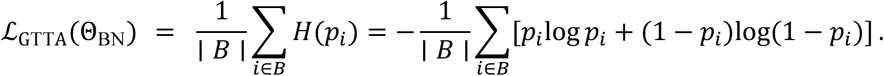

We took one adaptation step (*s* = 1) with Adam (learning rate 5 × 10^−5^; weight decay = 0). To prevent accumulation of batch-specific drift, we used an episodic update scheme in which Θ_BN_ was restored to its pre-adaptation values after each mini-batch. The adapted model provides a lightweight and fully label-free mechanism to reduce cohort–EHR distribution shift at test time.

### MedStitcher: Model training details

We trained and evaluated the model using a nested stratified cross-validation design with explicit separation of labeled source cohorts and auxiliary unlabeled target-domain data. Labeled patient nodes were drawn from PPMI and PDBP, whereas All of Us nodes were retained as unlabeled auxiliary nodes for graph construction and test-time adaptation only. In the outer evaluation loop, both the PPMI and PDBP labeled patient sets were independently partitioned into five stratified folds and then recombined fold-wise, such that each outer training and testing split preserved class balance within each cohort. To prevent information leakage, feature preprocessing was re-estimated within each outer fold: continuous variables were standardized using a StandardScaler fit on the outer training nodes only, and missing values were imputed using a constant-value imputer fit on the same training partition. The patient–patient similarity structure and the full heterogeneous graph were then rebuilt within each fold using the fold-specific preprocessed data. Hyperparameters were selected by a second five-fold stratified inner cross-validation performed within the outer training set, again preserving the PPMI/PDBP cohort structure during split construction. The candidate configurations varied residual connection use (True or False) and the number of graph attention heads (1 or 2), while other settings were fixed based on preliminary tuning: hidden dimension = 256, dropout = 0.2, number of graph layers = 3, batch normalization enabled, activation function = ReLU, learning rate = 1 × 10^−4^, and weight decay = 1 × 10^−5^. For each candidate setting, the patient/type-specific encoders and graph propagation module shared the same hidden dimensionality. The optimal configuration was selected according to mean validation AUROC across the inner folds. Model selection was based on the mean validation AUROC across inner folds. After selecting the best-performing configuration, the model was retrained on the complete outer training set for 50 epochs using the Adam optimizer. To account for class imbalance, the loss function was a weighted binary cross-entropy with logits, with the positive-class weight recalculated within each training split as the ratio of negative to positive samples. Mini-batch training was performed with neighborhood sampling, using up to three message-passing layers and a fixed neighbor budget per layer. During evaluation, predictions were generated with graph test-time adaptation, in which a lightweight entropy-minimization step updated a restricted subset of model parameters on unlabeled target batches, optionally supplemented by auxiliary All of Us nodes, before inference.

### Baseline models

#### XGBoost and LightGBM

As tabular machine-learning baselines, we implemented XGBoost^23^ and LightGBM^22^ using the same patient-level input features as the graph-based models, excluding identifiers and future-window variables. Specifically, all features were coerced to numeric format, and variables corresponding to later follow-up horizons were removed before model fitting. Training was performed in a nested cross-validation framework aligned with the PPMI/PDBP evaluation design. Within each outer split, missing values were imputed using a K-nearest-neighbors imputer fitted on the training partition only, followed by z-score standardization of continuous variables using statistics estimated from the same training set. To address class imbalance, synthetic minority over-sampling (SMOTE)^61^ was applied within the nested cross-validation pipeline. Hyperparameters were selected by inner cross-validation with area under the receiver operating characteristic curve (AUROC) as the primary selection criterion. For both XGBoost and LightGBM, the search space included the number of trees (*n*_*estimators* = 50 or 100) and learning rate (10^−3^ or 10^−2^). The final baseline predictions were generated from the best-performing model in each outer fold.

#### Conventional GNN

As a graph-based baseline without knowledge-graph augmentation, we constructed a patient-only graph in which node features consisted solely of tabular clinical and demographic attributes, without introducing external biomedical entities or knowledge-graph edges. In the initial graph construction, no feature nodes or knowledge-layer nodes were added. Instead, patient–patient connectivity was derived from feature-space similarity, with cosine similarity computed from the available patient attributes and used to define dynamic top-K (K = 40) nearest-neighbor edges during fold-specific graph construction. Thus, this baseline retained graph-based inductive bias through patient similarity propagation while excluding all explicit knowledge-graph information. Node representations were first mapped through a multilayer perceptron encoder and then passed to a graph neural network backbone with an edge-aware graph convolution/attention operator and a linear classification head. The model was trained using weighted binary cross-entropy with logits to account for class imbalance, and optimization was performed with Adam. Hyperparameters were tuned in nested cross-validation using AUROC on the validation folds, with the candidate settings including hidden dimension, dropout, number of graph propagation layers, batch normalization, residual connection, attention heads, learning rate and weight decay. This baseline therefore isolated the contribution of graph-based patient similarity modeling from the additional contribution of external knowledge-graph structure.

### Model interpretation using GNNExplainer^19^

*k* = 2PyTorch Geometric implementation of GNNExplainer in an edge-only setting, such that the explainer optimized an object-level edge mask. Explanations were generated separately for individual target patients on induced *k*-hop neighborhoods (*k* = 2) extracted from the full graph, which reduced memory burden and localized interpretation to the patient-specific decision context. Within each outer fold, seed patients were selected by grouping predictions into true positives, true negatives, false positives and false negatives using a probability threshold of 0.5. Within each group, patients were ranked by confidence, defined as the absolute distance between the predicted probability and the decision threshold, and up to five high-confidence cases per group were selected for patient-feature attribution. For graph-level explanation of the biomedical knowledge graph, representative seed patients were selected analogously from high-confidence positive-label, negative-label and misclassified cases, with cohort representation from both PPMI and PDBP preserved when possible.

For each selected seed patient, we extracted a patient-centered two-hop subgraph from the full heterogeneous graph and evaluated the frozen trained model on this subgraph. GNNExplainer was run for 120 optimization epochs with node-level binary-classification output defined on the raw logits. To derive biologically interpretable graph motifs, we filtered each patient-specific explanation to retain only non-patient biomedical knowledge graph edges, thereby excluding patient nodes and preserving relations among biomedical entities. For each explained patient, the top 20 non-patient biomedical knowledge graph edges ranked by GNNExplainer edge-importance score were retained. To estimate the direction and magnitude of each retained relation’s contribution to the target prediction, we performed perturbation analysis by removing each retained edge in turn through zeroing its edge-feature row and recomputing the change in the target patient logit.

Patient-specific biomedical knowledge graph explanation subgraphs were aggregated within each outer fold by canonicalizing undirected edges, summing perturbation-based edge contributions across selected patients and recording edge support as the number of patient-specific explanation subgraphs in which each edge appeared. Fold-level biomedical knowledge graph explanation subgraphs retained edges with support ≥1 and were capped at the top 200 aggregated knowledge-graph-only edges ranked by absolute signed contribution. The “core stitcher” subgraph shown in Fig. 3b was defined from these retained biomedical knowledge graph relations and their incident nodes. Thus, node inclusion in the core stitcher was determined by incidence to retained explanatory edges, and edge support was reported as a descriptive measure of recurrence across selected patient-specific explanations rather than as an additional high-support cutoff beyond support ≥1.

#### Patient-level feature importance

Patient-level feature attribution was computed separately from graph-structure interpretation using an integrated-gradients-style procedure applied to the trained patient encoder and graph model on patient-centered two-hop subgraphs. For each selected outer-test patient, we constructed a subgraph wrapper that returned only the logit of the target patient node and computed feature attributions relative to a zero baseline using 16 interpolation steps. Only the attribution vector of the target patient node was retained. Features with no variation across patient nodes were excluded before aggregation. For each explained patient, features were ranked by the absolute value of their signed attribution, and the top 20 features were retained. Global patient-feature importance was summarized by aggregating mean absolute attributions across selected representative patients from all outer folds, rather than across all outer-test patients. To visualize the distribution of feature effects, we generated SHAP-like beeswarm plots in which each point represented one patient-specific signed attribution value for a given feature; features were ordered by mean absolute attribution, and positive versus negative effects were displayed according to the sign of the attribution.

### Evaluation of MedStitcher-predicted probable rapid progressors in RWD through survival analysis

In RWD, we measured PD progression using onset of cognitive impairment, dementia, and fall events as advanced outcomes. We examined time to first recorded event using Kaplan–Meier (KM) estimators and compared risk strata with two-sided log-rank tests. For falls, which can recur over follow-up, we further used a recurrent-event Cox proportional hazards model under the Andersen–Gill counting-process formulation.

We retrained MedStitcher over entire datasets using hyperparameters with the best performance. Using the model, we predicted a risk score for each PD patient in the All-of-Us dataset, indicating the estimated possibility that this patient belongs to the fast progressor. We then stratified all All-of-Us PD patients into rapid (top 20%) and non-rapid (remaining 80%) groups.

Since lack of PD progression subtype label and clinical assessments measuring PD progression speed, we used onset of cognitive impairment, dementia, and fall events as advanced outcomes. We examined time to first recorded event using Kaplan–Meier (KM) estimators, comparing risk strata between the rapid and non-rapid progressor groups, using two-sided log-rank tests. For falls, which can recur over follow-up, we further used a recurrent-event Cox proportional hazards model under the Andersen–Gill counting-process formulation. Specifically, we defined the same model-based risk strata and partitioned each patient’s follow-up into successive intervals [*t_start_*, *t_stop_*) bounded by observed fall events, with administrative censoring at the earlier of 10 years or 2025-07-31. We then fit an adjusted Andersen–Gill Cox model adjusted by age, sex, race, and ethnicity to account for within-person correlation across recurrent events.

### Identification and validation of early indicators of rapid progression in PD

We further tested whether the model extracts transferable clinical signals at the phenotype level across independent EHR systems. Diagnosis codes in All-of-Us and INSIGHT were mapped to phecodes using established ICD-to-phecode mappings (phecode v1.2 / PheWAS resources) to enable clinically meaningful aggregation of EHR diagnoses^62,63^. We include all phecodes^62^ observed at least once (1732 phecodes). For each phecode, we fitted a logistic regression model to evaluate its association with rapid PD progression while adjusting for age, sex, race, and ethnicity. Multiple testing was controlled using Bonferroni correction (two-sided; significance threshold P < 0.05/1732).

To assess cross-system reproducibility, phecodes significantly associated with rapid progression in All-of-Us were further evaluated in an independent large-scale real-world dataset, the INSIGHT database. A PD cohort was first constructed in INSIGHT using the same inclusion criteria applied in All-of-Us. For each phecode, patients were classified as carriers or non-carriers based on diagnoses recorded within the same baseline observation window. We then fitted Cox proportional hazards models to test differences in PD progression rates, defined as time to the incidence of PD-related outcomes including dementia, cognitive impairment, and falls. Models were adjusted for age, sex, race, and ethnicity. Hazard ratios with 95% confidence intervals were estimated over a 10-year follow-up period to quantify the reproducibility of phenotype-level risk signals across independent EHR systems.

### Data visualization

We used UMAP embedding^24^ for visualization of patient and knowledge graph entities in 2D embedding space. Network visualization was done using Cytoscape^64^.

### Pathway Enrichment

The top 30 gene nodes ranked by node importance weight in the core stitcher network were used for pathway enrichment analysis with g:Profiler^65^. Enrichment was assessed using KEGG^66^, Reactome^67^, and WikiPathways^68^ annotations, with the annotated gene set as the statistical background. Enrichment P values were adjusted for multiple testing, and pathways with an adjusted P value < 0.05 were considered significant. Significant pathways were manually grouped according to their biological functions.

## Data Availability

This study used data from the Parkinson's Progression Markers Initiative PPMI at www.ppmi-info.org/access-data-specimens/download-data, the Parkinson Disease Biomarkers Program PDBP at https://pdbp.ninds.nih.gov, the AMP-PD Knowledge Platform at http://amp-pd.org, the All of Us Research Program Controlled Tier Dataset V8 through the All of Us Researcher Workbench at https://researchallofus.org, and the INSIGHT Clinical Research Network. Information about INSIGHT is available at https://insightcrn.org. These datasets are available to qualified or authorized users subject to their respective data access procedures, data use agreements, and institutional approvals. Individual-level participant data cannot be redistributed by the authors.

## Data availability

This study used data from PPMI (www.ppmi-info.org/access-data-specimens/download-data, RRID:SCR 006431) and PDBP (https://pdbp.ninds.nih.gov). Data from the PDBP were download from AMP-PD Knowledge Platform (http://amp-pd.org). This study used data from the All of Us Research Program’s Controlled Tier Dataset V8, available to authorized users on the All of Us Researcher Workbench (https://researchallofus.org). Information of the INSIGHT database is available at https://insightcrn.org.

## Code availability

Computer codes for this study are available at https://github.com/pkuyzy/MedStitcher.

## Acknowledgements

This study used data from the PPMI (obtained from PPMI database at www.ppmi-info.org/access-data-specimens/download-data) and PDBP (obtained through AMP-PD Knowledge Platform at https://www.amp-pd.org). PPMI – a public-private partnership – is funded by the Michael J. Fox Foundation for Parkinson’s Research and funding partners, including 4D Pharma, Abbvie, AcureX, Allergan, Amathus Therapeutics, Aligning Science Across Parkinson’s, AskBio, Avid Radiopharmaceuticals, BIAL, BioArctic, Biogen, Biohaven, BioLegend, BlueRock Therapeutics, Bristol-Myers Squibb, Calico Labs, Capsida Biotherapeutics, Celgene, Cerevel Therapeutics, Coave Therapeutics, DaCapo Brainscience, Denali, Edmond J. Safra Foundation, Eli Lilly, Gain Therapeutics, GE HealthCare, Genentech, GlaxoSmithKline plc (GSK), Golub Capital, Handl Therapeutics, Insitro, Janssen Neuroscience, Jazz Pharmaceuticals, Lundbeck, Merck, Meso Scale Discovery, Mission Therapeutics, Neurocrine Biosciences, Neuropore, Pfizer, Piramal, Prevail Therapeutics, Roche, Sanofi, Servier, Sun Pharma Advanced Research Company, Takeda, Teva, UCB, Vanqua Bio, Verily, Voyager Therapeutics, the Weston Family Foundation and Yumanity Therapeutics. For up-to-date information on the study, visit www.ppmi-info.org. PDBP is supported by the National Institute of Neurological Disorders and Stroke at the National Institutes of Health. Investigators include: Roger Albin, Roy Alcalay, Alberto Ascherio, Thomas Beach, Sarah Berman, Bradley Boeve, F. DuBois Bowman, Shu Chen, Alice Chen-Plotkin, William Dauer, Ted Dawson, Paula Desplats, Richard Dewey, Ray Dorsey, Jori Fleisher, Kirk Frey, Douglas Galasko, James Galvin, Dwight German, Lawrence Honig, Xuemei Huang, David Irwin, Kejal Kantarci, Anumantha Kanthasamy, Daniel Kaufer, James Leverenz, Carol Lippa, Irene Litvan, Oscar Lopez, Jian Ma, Lara Mangravite, Karen Marder, Laurie Orzelius, Vladislav Petyuk, Judith Potashkin, Liana Rosenthal, Rachel Saunders-Pullman, Clemens Scherzer, Michael Schwarzschild, Tanya Simuni, Andrew Singleton, David Standaert, Debby Tsuang, David Vaillancourt, David Walt, Andrew West, Cyrus Zabetian, Jing Zhang, and Wenquan Zou. The PDBP Investigators have not participated in reviewing the data analysis or content of the manuscript. For up-to-date information on the study, visit: https://pdbp.ninds.nih.gov. The AMP-PD program is a public-private partnership managed by the Foundation for the National Institutes of Health and funded by the National Institute of Neurological Disorders and Stroke in partnership with the Aligning Science Across Parkinson’s initiative, Celgene, GSK, the Michael J. Fox Foundation for Parkinson’s Research, Pfizer, AbbVie, Sanofi, and Verily Life Sciences. For up-to-date information on the study, visit https://www.amp-pd.org.

We gratefully acknowledge All of Us participants for their contributions, without whom this research would not have been possible. We also thank the National Institutes of Health’s All of Us Research Program for making available the participant data examined in this study.

This work was supported by the following: The Michael J. Fox Foundation for Parkinson’s Research to F.W. and C.S.; and the National Institute of Neurological Disorders and Stroke of the National Institutes of Health to C.S. (R01NS140142 and R01NS148533).

## Author contributions

F.W. and C.S. conceived the study. F.W. and C.S. secured funding and supervised the research. F.W., C.S., and Z.Y. developed MedStitcher. Z.Y. and H.L. cleaned clinical data. Z.Y., H.L. and M.Z. performed omics data processing. Z.Y. and Z.H. processed biomedical knowledge graph (BKG) and connected to patient data. Z.Y. led model deployment and data analysis. W-T.W. and S.J. performed data visualization. C.S. and Z.Y. drafted manuscript. All authors critically revised the manuscript for important intellectual content. All authors provided comments and approved the paper.

## Extended Data Figures

**Extended Data Fig. 1.**
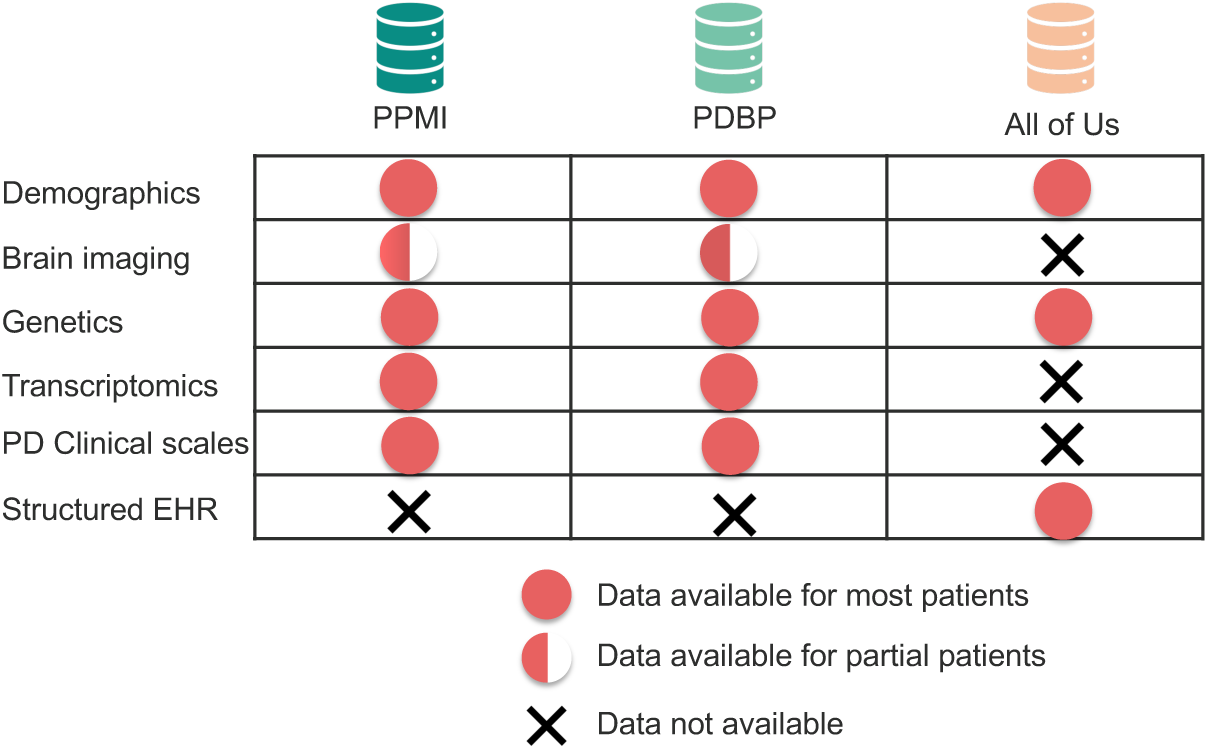
Modality availability of PPMI and PDBP (PD research cohort studies) and All of Us (real-world data).

**Extended Data Fig. 2.**
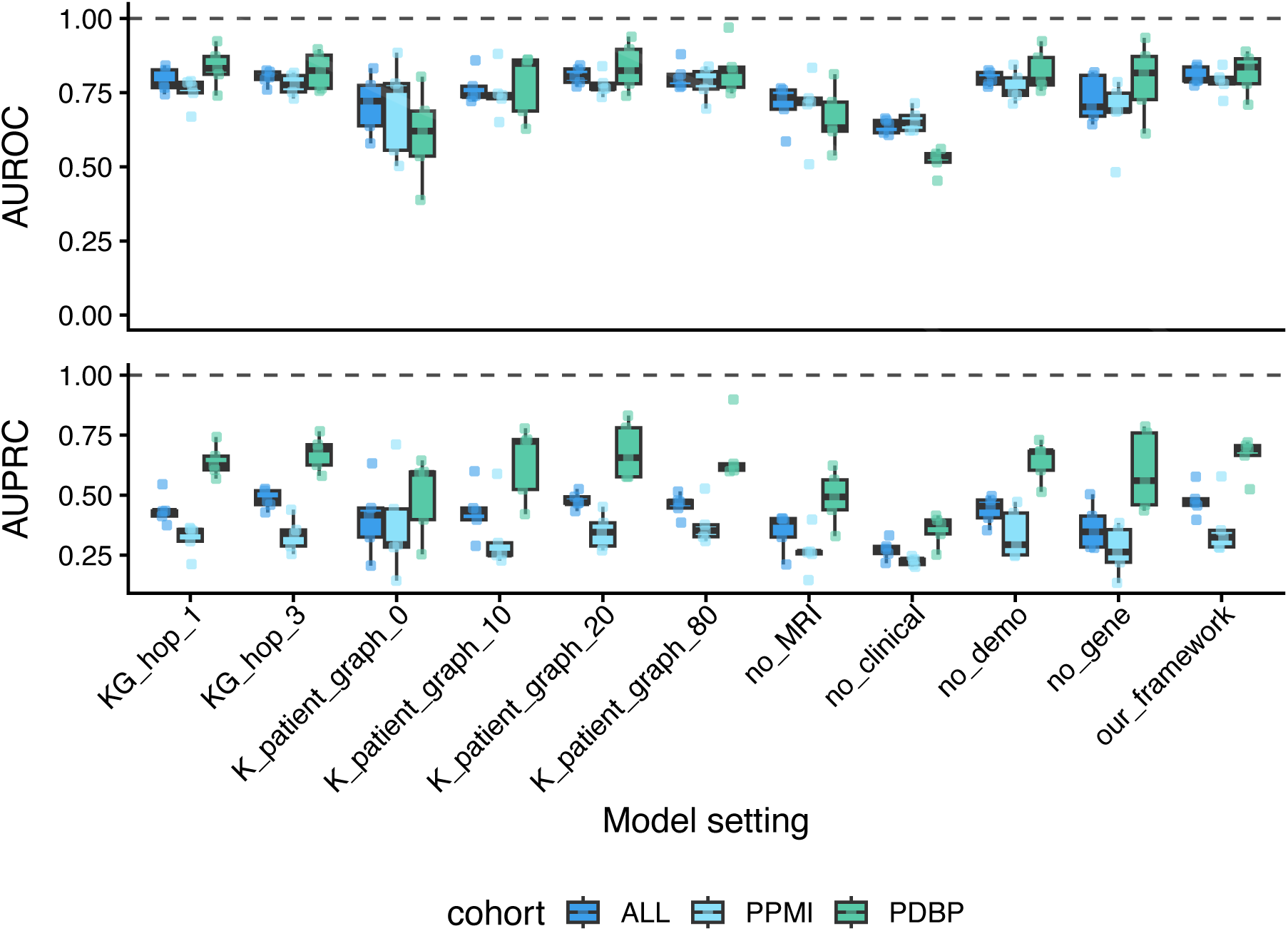
Ablation and robustness analyses of knowledge-graph integration and modality contributions across cohorts. Performance comparison of alternative model settings evaluated in labeled research cohorts (PPMI and PDBP) and pooled analysis (ALL). Upper, AUROC; lower, AUPRC. Box plots summarize five-fold outer cross-validation results, with points indicating fold-specific performance. Model settings include varying knowledge-graph (KG) neighborhood depth (KG_hop_1, KG_hop_3), patient–patient graph construction (K_patient_graph with K = 0, 10, 20, 80), modality ablations (no_MRI, no_clinical, no_demo, no_gene), and the full framework (“our_framework”). Increasing KG neighborhood depth and incorporating patient similarity edges consistently improved discrimination relative to shallow or disconnected configurations, supporting the contribution of structured biomedical context. Removal of key modalities—particularly clinical assessments and genetic information—led to marked reductions in performance, highlighting their complementary roles in progression modeling. Across both AUROC and AUPRC, the full framework achieved the most stable and highest overall performance in pooled and cohort-specific evaluations, demonstrating robustness to cohort heterogeneity and reinforcing the importance of knowledge-graph augmentation and multimodal integration.

**Extended Data Fig. 3.**
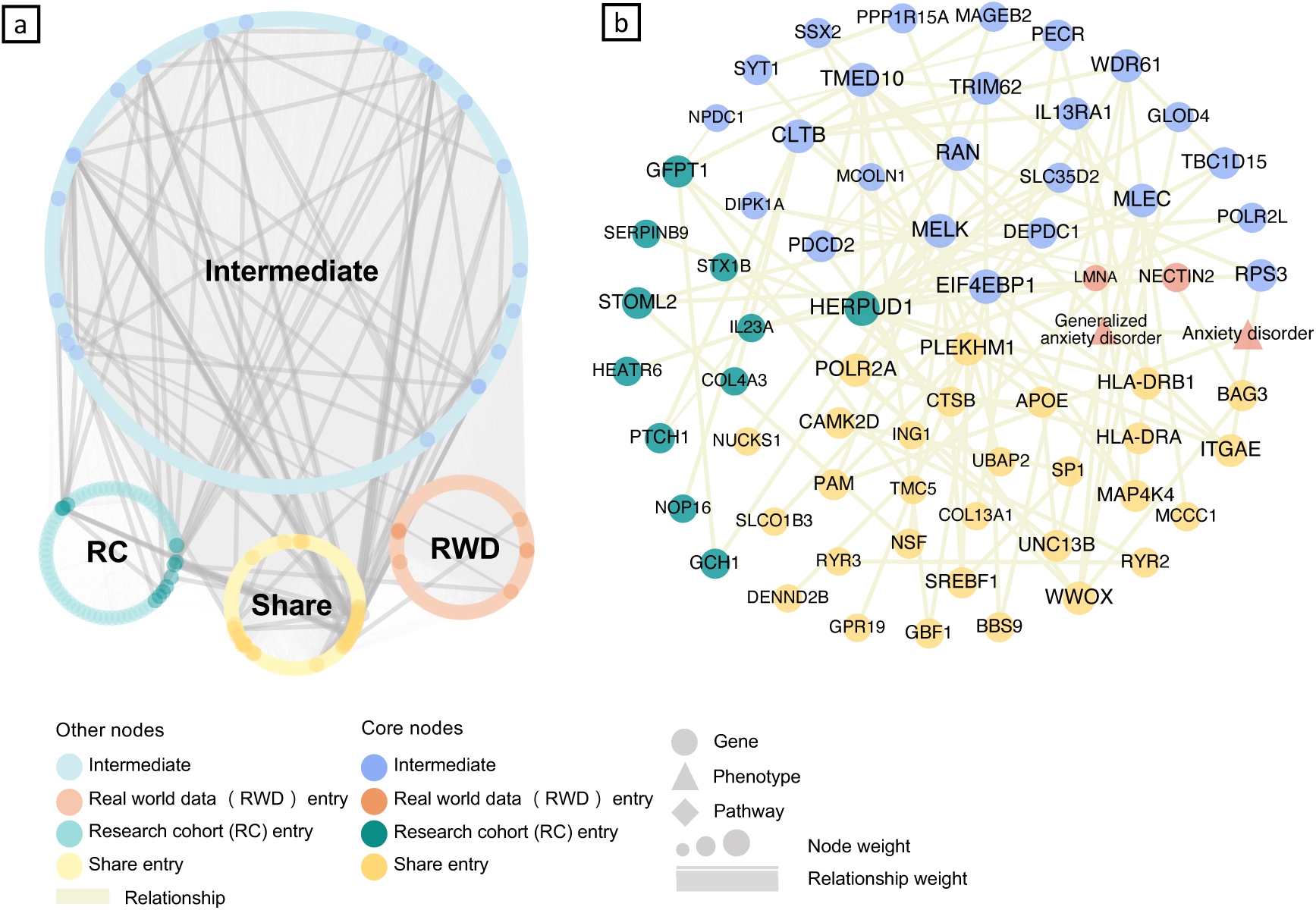
Important subgraph of the BKG in predicting rapid progressors in All of Us data. **a,** Core-stitcher subgraph within the BKG, showing intermediate nodes that bridge research-cohort (RC), real-world data (RWD) and shared entry nodes. **b,** Entity-level view of the core stitcher, showing gene, phenotype and pathway nodes; node size and edge width represent explanation weight and relationship weight, respectively.

