## Supplementary Material for "Early Prediction of Parkinson’s Disease Progression by Integrating Research Cohort and Real-World Data Using Knowledge-Anchored Graph Learning"

**Supplementary Note 1.** Theoretical analysis on why KG improves PD-progression prediction

In the main text, we empirically show that augmenting the patient graph with a KG improves prediction of rapid PD progression. Here, we provide a theoretical rationale for why KG augmentation can be beneficial under real-world modality incompleteness. In routine clinical and multi-omics settings, patient records are inherently heterogeneous and partially observed: different cohorts or health systems capture different subsets of clinical assessments, molecular measurements and imaging features, and missingness patterns are often cohort-specific. When key modalities are unobserved, patients with distinct progression trajectories may become less distinguishable using measured features alone.

Intuitively, KG augmentation introduces structured biomedical entities and relations that provide mechanistic context and can act as proxy information for unmeasured or sparsely measured modalities. Message passing over the KG-augmented topology allows the model to propagate and aggregate disease-relevant signals through biologically grounded connections, enriching patient representations beyond directly observed features. This added context can increase the separability of patients with different progression trajectories under the same observed data, offering an explanation for the empirical gains reported in the main text.

Below, we provide a formal proof on why KG augmentation can be beneficial under real-world modality incompleteness.

**Problem setup**

Let $\mathcal{M} = \{m_1 \dots m_K\}$ denote data modalities (e.g., clinical records, genetics). For a patient $u$, let $X_m(u)$ be the feature from modality $m$. Real datasets exhibit **modality missingness**: only a subset $S \subseteq M$ is observed. Write $X_S(u) = \{X(u): m \in S\}$.

Let $G_s$ be the **patient graph** derived from observed modalities $S$ (nodes, edges, and attributes constructed from $X_s$); we view the graph construction as fixed given $X_s$. The binary label $y(u) \in \{0,1\}$ indicates PD rapid progression for patient $u$.

Let $\mathcal{H}$ be a family of real-valued graph functions on $\mathcal{G}_S$ $h: G_s \to \mathbb{R}$ and let $\mathcal{N}$ be a family of multilayer perceptrons (MLPs). We consider a "feature extractor + MLP head" hypothesis family

$$H_{GNN} = \{\mathrm{h}_{\mathcal{N}} \circ \phi : \phi(\mathrm{G}) = \left(\mathrm{f}_1(\mathrm{G}), \dots, \mathrm{f}_\mathrm{d}(G)\right),\ \mathrm{d} \in \mathcal{N},\ \mathrm{f}_\mathrm{i} \in \mathcal{H},\ h_{\mathcal{N}} \in \mathcal{N}\},$$

where the final prediction can be thresholded to $\in \{0,1\}$ when needed. This abstraction captures "GNN feature extractor + shallow MLP head".

Let $KG$ be a fixed biomedical knowledge graph containing clinical entities and relations (e.g., diseases–genes–symptoms). From $G_s$ and $KG$, construct an **augmented graph** $\tilde{G}_{S,\mathrm{KG}} = (G_s, KG)$**,** by adding: (i) KG entity nodes referenced by concepts present/derivable from $X_s$ ; (ii) edges among those entities as in $KG$; and (iii) cross-links from patient (or patient-feature) nodes to the attached $KG$ entities (e.g.,symptom/disease links, gene–disease edges). Let $H_{GNN+KG}(S)$ be the analogous hypothesis family operating on $\tilde{G}_{S,\mathrm{KG}}$. We now define **expressiveness (task-relative)**:

***Definition 1. (Task expressiveness under observed modalities; realizable setting).***

Assume a deterministic target function $y: G_s \to \{0, 1\}$ on the domain $G_s$. We say that a function class $\mathcal{H}$ **distinguishes labels** on $\mathcal{G}_S$ $\left(same\ for\ \tilde{G}_{S,\mathrm{KG}}\right)$ if

$$\forall\, G^{[1]}, G^{[2]} \in \mathcal{G}_S \text{ with } y\left(G^{[1]}\right) \neq y\left(G^{[2]}\right), \exists\, f \in \mathcal{H} \text{ s.t. } f\left(G^{[1]}\right) \neq f\left(G^{[2]}\right).$$

Equivalently, $\mathcal{H}$ separates every label-discordant pair in a pairwise sense.

**Finite-domain bridge (discrimination ⇔realizability)**

***Lemma 1 (From pairwise discrimination to zero error on finite domains)***

Suppose $\mathcal{G}_S$ is finite and $\mathcal{H}$ distinguishes labels on $\mathcal{G}_S$. Then there exists $h \in \mathcal{H}_{GNN}$ such that $h(G) = y(G)$ for all $G \in \mathcal{G}_S$ (that is, zero population error on $\mathcal{G}_S$).

*Proof.* Because $\mathcal{G}_S$ is finite, the set of label-discordant pairs is finite. For each discordant pair $(G^i, G^j)$ with $y\left(G^i\right) \neq y\left(G^j\right)$, choose one function $f_{ij} \in \mathcal{H}$ such that $f_{ij}\left(G^i\right) \neq f_{ij}\left(G^j\right)$. Let $\phi(G) \in \mathbb{R}^d$ be the vector collecting all such $f_{ij}(G)$, where $d$ is the number of discordant pairs. Then for any $G^i, G^j with$ different labels, the coordinate corresponding to $(i, j)$ differs, hence $\phi(G_i) \neq \phi\left(G_j\right)$.

Define the embedded sets $\Phi_0 = \{\phi(G) : y(G) = 0\}\ and\ \Phi_1 = \{\phi(G) : y(G) = 1\}$, which are disjoint finite subsets of $\mathbb{R}^d$. Because $\Phi_0$ and $\Phi_1$ are disjoint, there exists a continuous function $g: \mathbb{R}^d \to \mathbb{R}$ such that $g(x) > 0$ for $x \in \Phi_1$ and $g(x) < 0$ for $x \in \Phi_0$ (for example, one can construct $g$ using distance-to-set functions). By universal approximation results for MLPs (for example, deep ReLU networks), there exists $h_{\mathcal{N}} \in \mathcal{N}$ that approximates $g$ sufficiently well on the finite set $\Phi_0 \cup \Phi_1$ so that $\mathrm{sign}(h_{\mathcal{N}}(x)) = \mathrm{sign}(g(x))$ on that set[1]. Therefore $h(G) = \mathbf{1}\{h_{\mathcal{N}}(\phi(G)) > 0\}$ satisfies $h(G) = y(G)$ for all $G \in \mathcal{G}_S$. ■

***Lemma 2 (Converse: realizability implies pairwise discrimination)***

If there exists $h \in \mathcal{H}_{\text{GNN}}$ such that $h(G) = y(G)$ for all $G \in \mathcal{G}_S$, then $\mathcal{H}$ distinguishes labels on $\mathcal{G}_S$.

*Proof.* Let $G^{[1]}, G^{[2]}$ be any pair with $y(G^{[1]}) \neq y(G^{[2]})$. Since $h(G) = y(G)$, we have $h(G^{[1]}) \neq h(G^{[2]})$. But $h = h_{\mathcal{N}} \circ (f_1, \ldots, f_d)$ for some $f_i \in \mathcal{H}$. If all $f_i(G^{[1]}) = f_i(G^{[2]})$, then the inputs to $h_{\mathcal{N}}$ would be identical and the outputs could not differ. Hence there exists at least one coordinate $f_j \in \mathcal{H}$ such that $f_j(G^{[1]}) \neq f_j(G^{[2]})$. ■

***Proposition 1 (Equivalence on finite $\mathcal{G}_S$)***

When $\mathcal{G}_S$ is finite, the following are equivalent: (i) $\mathcal{H}$ distinguishes labels on $\mathcal{G}_S$; (ii) there exists $h \in \mathcal{H}_{\text{GNN}}$ achieving zero error on $\mathcal{G}_S$.

*Proof.* (i)⇒(ii) follows from Lemma 1; (ii)⇒(i) follows from Lemma 2. ■

This definition and the two lemmas mirror how GNN expressivity is treated in the literature (hypothesis class capacity on a given input domain), while acknowledging that in healthcare the input domain itself changes when a KG is attached[2–4].

We then prove the **expressivity gain from KG augmentation under modality missingness.**

**Setup.**

Let $S \subseteq \mathcal{M}$ denote the set of observed modalities. For each patient instance $u$, let $G_S(u)$ be the patient graph constructed from observed features $X_S(u)$. Define a deterministic augmentation operator

$$\mathcal{A}_{\text{KG}}: G_S \mapsto \tilde{G}_{S,\text{KG}}$$

that attaches (i) KG entity nodes grounded in concepts present/derivable from $X_S$, (ii) KG edges among attached entities, and (iii) cross-links between patient (or patient-feature) nodes and KG entities. Let $\pi$ be the projection that removes all KG-added nodes/edges:

$$\pi(\tilde{G}_{S,\mathrm{KG}}) = G_S.$$

We consider message-passing GNN families with permutation-invariant aggregation and a permutation-invariant readout (the standard setting in expressivity analyses[2–4]). Let $H_{\mathrm{GNN}}(S)$ be the hypothesis class induced by applying a chosen GNN family to graphs $G_S$. Let $H_{\mathrm{GNN+KG}}(S)$ be the analogous class when the same family is applied to augmented graphs $\tilde{G}_{S,\mathrm{KG}}$.

**Assumption (edge-type controllability).** All KG-incident edges (patient–KG links and KG–KG edges) are represented as one or more designated relation/edge types, and the GNN family contains parameterizations that can null out messages carried by selected edge types (e.g., relation-specific weights, gates, or attention scores that can be set to zero). This is satisfied by common typed-edge message passing models (e.g., R-GCN–style or attention with relation/gate parameters).

***Theorem 1 (Monotonicity under KG augmentation).***

For any observed modality set $S$,

$$\{h \circ \pi : h \in H_{GNN}(S)\} \subseteq H_{GNN+KG}(S).$$

*Proof.*

Fix any $h \in H_{\mathrm{GNN}}(S)$. Consider the same GNN architecture applied to $\tilde{G}_{S,\mathrm{KG}}$, with identical parameters on the original edge types and node features, but with all parameters (weights/gates/attention) associated with KG-incident edge types set to zero. Then no message can pass through KG-added edges, so the computation on $\tilde{G}_{S,\mathrm{KG}}$ reduces exactly to the computation on $\pi(\tilde{G}_{S,\mathrm{KG}}) = G_S$. Hence this augmented-graph model realizes $h \circ \pi$, proving the inclusion. ■

*Remark.* Theorem 1 is a pure hypothesis-class statement: adding KG cannot reduce what the model family can represent, because the model can always ignore the KG.

***Theorem 2 (Strict gain under missing-modality dependence)***

Assume there exist two modalities $m_1 \notin S$(unobserved) and $m_2 \in S$(observed), such that the label $Y$ depends on both, e.g.,

$$Y = f(X_{m_1}, X_{m_2}),$$

and the following hold:

1. (**Observed-graph indistinguishability**) There exist two patient instances $u, v$ such that $G_S(u) \cong G_S(v)$(equivalent for GNN) but $Y(u) \neq Y(v)$. In particular, any predictor measurable w.r.t. $G_S$ cannot perfectly separate $u$ and $v$.
2. (**KG reveals a proxy of the missing modality**) The biomedical KG exposes a deterministic or high-fidelity link from signals derivable from $X_{m_2}$ to a proxy of $X_{m_1}$ via short clinical paths (e.g., symptom→disease, disease→gene, gene→pathway) so that $\tilde{G}_{S,\mathrm{KG}}(u)$ and $\tilde{G}_{S,\mathrm{KG}}(v)$ are not equivalent for GNN, and this difference correlates with $Y$.

Then

$$\mathcal{H}_{\mathrm{GNN}}(S) \subset \mathcal{H}_{\mathrm{GNN+KG}}(S)$$

*Proof.*

**Existence of the $\mathbf{G_S(u)}$, $\mathbf{G_S(v)}$, $\mathbf{\tilde{G}_{S,KG}(u)}$, and $\mathbf{\tilde{G}_{S,KG}(v)}$.** An example is shown in Supplementary Fig. 1. We will prove that the two conditions hold.

**Indistinguishability w.r.t. observed graph**. Formally, Let $V = P \cup M$ with $P = \{p_1, p_2, p_3, p_4\}$ (patient nodes) and $M = \{m_1, m_2, m_3, m_4\}$ (modality/KG nodes). Assume **type-homogeneous features**:

$$h_p^{(0)} = x_P(\forall p \in P), h_m^{(0)} = x_M(\forall m \in M),$$

where $x_P, x_M \in \mathbb{R}^{d_0}$are fixed.

Let $G^{(a)} = \left(V, E^{(a)}\right)$and $G^{(b)} = \left(V, E^{(b)}\right)$ denote the two graphs in Supplementary Fig. 1 (a) and (b):

- (a) two disjoint length-3 paths:

$$(m_1, p_1, p_3, m_3), (m_2, p_2, p_4, m_4).$$

- (b) two disjoint length-3 paths:

$$(m_1, p_1, p_2, m_2), (m_3, p_3, p_4, m_4).$$

Edges are undirected and unlabelled (edge features, if any, are constant and identical across graphs).

A GNN layer has the form[3]:

$$h_v^{(t+1)} = \mathrm{AGG}_{u \in \mathcal{N}(v)} \phi^{(t)} \left(h_v^{(t)}, h_u^{(t)}\right)$$

where $AGG$ is **permutation-invariant** (sum/mean/max or attention-weighted sum with shared parameters), and $\phi^{(t)}, \psi^{(t)}$are shared across all nodes/edges. We note that the definition can be used for all message-passing-based GNNs[3], where their expressiveness is bounded by the so-called 1st order Weisfeiler-Leman algorithm[5].

We let $h_v^{(t)}$denote the hidden states on $G^{(a)}$and $\tilde{h}_v^{(t)}$those on $G^{(b)}$when running the same GNN with the same parameters.

Define a bijection $\pi: V \rightarrow V$ by

$$\pi(m_1) = m_1, \pi(p_1) = p_1, \pi(p_3) = p_2, \pi(m_3) = m_2, \pi(m_2) = m_3, \pi(p_2) = p_3, \pi(p_4) = p_4, \pi(m_4) = m_4.$$

One checks directly that $(u, v) \in E^{(a)} \iff (\pi(u), \pi(v)) \in E^{(b)}$ and that $\pi$ **preserves types** $(P \to P,\ M \to M)$. Hence $G^{(b)} \cong G^{(a)}$ via $\pi$.

We prove by induction on $t$ that

$$\boxed{\tilde{h}^{(t)}_{\pi(v)} = h^{(t)}_v \text{ for all } v \in V} \qquad (*)$$

**Base ($t = 0$).** By type-homogeneous initialization and the fact that $\pi$ preserves types, $\tilde{h}^{(0)}_{\pi(v)} = h^{(0)}_v$ for all $v$.

**Inductive step.** Assume $(*)$ holds at depth $t$. Fix any $v \in V$. Because $\pi$ is an isomorphism,

$$\mathcal{N}_{G^{(b)}}(\pi(v)) = \{\pi(u) : u \in \mathcal{N}_{G^{(a)}}(v)\}.$$

Hence, using parameter sharing, the inductive hypothesis, and permutation-invariance of AGG,

$$\begin{aligned}
\tilde{h}^{(t+1)}_{\pi(v)} &= \mathrm{AGG}_{w \in \mathcal{N}_{G^{(b)}}(\pi(v))} \phi^{(t)}\left(\tilde{h}^{(t)}_{\pi(v)}, \tilde{h}^{(t)}_w\right) \\
&= \mathrm{AGG}_{u \in \mathcal{N}_{G^{(a)}}(v)} \phi^{(t)}\left(\tilde{h}^{(t)}_{\pi(v)}, \tilde{h}^{(t)}_{\pi(u)}\right) \\
&= \mathrm{AGG}_{u \in \mathcal{N}_{G^{(a)}}(v)} \phi^{(t)}\left(h^{(t)}_v, h^{(t)}_u\right) = h^{(t+1)}_v.
\end{aligned}$$

Thus $(*)$ holds for $t + 1$. By induction, $(*)$ is true for all layers $t = 0, 1, \ldots, T$. Therefore for node-wise output, if the final predictor is a shared head $o_v = \ell\left(h^{(T)}_v\right)$, then

$$\tilde{o}_{\pi(v)} = \ell\left(\tilde{h}^{(T)}_{\pi(v)}\right) = \ell\left(h^{(T)}_v\right) = o_v,$$

so patient-node predictions match one-to-one via $\pi$.

In terms of graph-level output, any permutation-invariant readout $y = \rho\left(h_v^{(T)} : v \in V\right)$(sum/mean/max, attention-pooling with shared parameters, etc.),

$$\tilde{y} = \rho\left(\tilde{h}_w^{(T)} : w \in V\right) = \rho\left(h_v^{(T)} : v \in V\right) = y,$$

because $\tilde{h}_w^{(T)} : w \in V = h_v^{(T)} : v \in V$as multisets by $(*)$.

**KG makes the latent dependence observable.** We show that, for the two graphs in Supplementary Figure 6(c) and (d) with the orange KG edge $(m_1, m_2)$, there exists a choice of $\phi^{(t)}$and depth $T = 3$such that at least the patient embeddings—and hence any injective readout—are different across (a) and (b).

*Setting and assumptions*

- Nodes: $P = \{p_1, p_2, p_3, p_4\}$(patients), $M = \{m_1, m_2, m_3, m_4\}$ (KG/modality).
- Initial features (type-homogeneous): $h_p^{(0)} = x_P$for all $p \in P$, and $h_m^{(0)} = x_M$for all $m \in M$.
- Edges (undirected, no edge features):
  - (a): $(m_1, p_1), (p_1, p_3), (p_3, m_3)$and $(m_2, p_2), (p_2, p_4), (p_4, m_4)$plus $(m_1, m_2)$.
  - (b): $(m_1, p_1), (p_1, p_2), (p_2, m_2)$and $(m_3, p_3), (p_3, p_4), (p_4, m_4)$plus $(m_1, m_2)$.

*Layer-by-layer separation (three rounds)*

We compute only **types** of hidden states that will suffice to separate the graphs.

*Round 1*

For modality nodes:

$$
\begin{aligned}
h_{m_1}^{(1)} &= \phi^{(0)}(x_M, x_P) + \phi^{(0)}(x_M, x_M), \\
h_{m_2}^{(1)} &= \phi^{(0)}(x_M, x_P) + \phi^{(0)}(x_M, x_M), \\
h_{m_3}^{(1)} &= \phi^{(0)}(x_M, x_P), h_{m_4}^{(1)} = \phi^{(0)}(x_M, x_P).
\end{aligned}
$$

Thus the modalities split into two distinct states:

$$M^{\uparrow} \coloneqq h_{m_1}^{(1)} = h_{m_2}^{(1)} \neq M^{\downarrow} \coloneqq h_{m_3}^{(1)} = h_{m_4}^{(1)}.$$

Every patient has one patient and one modality neighbor at $t = 0$, so for all $p \in P$,

$$h_p^{(1)} = \phi^{(0)}(x_P, x_P) + \phi^{(0)}(x_P, x_M) =: P^{(1)}.$$

*Round 2*

Patients now see one modality in $M^{\uparrow}, M^{\downarrow}$and one patient in state $P^{(1)}$; hence they split into two distinct states:

$$
\begin{aligned}
P^{\uparrow} &\coloneqq \phi^{(1)}\left(P^{(1)}, M^{\uparrow}\right) + \phi^{(1)}\left(P^{(1)}, P^{(1)}\right) = h_{p_1}^{(2)} = h_{p_2}^{(2)}, \\
P^{\downarrow} &\coloneqq \phi^{(1)}\left(P^{(1)}, M^{\downarrow}\right) + \phi^{(1)}\left(P^{(1)}, P^{(1)}\right) = h_{p_3}^{(2)} = h_{p_4}^{(2)}.
\end{aligned}
$$

By injectivity and $M^{\uparrow} \neq M^{\downarrow}$, we have $P^{\uparrow} \neq P^{\downarrow}$.

Modalities remain in their two classes (their neighbor states differ but in the same way across (c) and (d)), so up to depth 2 the multiset of node states is identical between (c) and (d).

*Round 3 (the separating round)*

Now the graphs differ in how patients connect to patients:

- In (c): $p_1$(which is in class $P^{\uparrow}$) is adjacent to $p_3$(class $P^{\downarrow}$).
  Hence

$$h_{p_1}^{(3)} = \phi^{(2)}(P^{\uparrow}, M^{\uparrow}) + \phi^{(2)}(P^{\uparrow}, P^{\downarrow}).$$

  Symmetrically for $p_2$(with $p_4$) and for the down-class patients.

- In (b): $p_1$(class $P^{\uparrow}$) is adjacent to $p_2$(also $P^{\uparrow}$).
  Hence

$$\tilde{h}_{p_1}^{(3)} = \phi^{(2)}(P^{\uparrow}, M^{\uparrow}) + \phi^{(2)}(P^{\uparrow}, P^{\uparrow}).$$

Because $P^{\uparrow} \neq P^{\downarrow}$and the multiset encoder

$$\mathcal{M} \mapsto \sum_{z \in \mathcal{M}} \phi^{(2)}(P^{\uparrow}, z)$$

is **injective**, we obtain

$$h_{p_1}^{(3)} \neq \tilde{h}_{p_1}^{(3)}.$$

By the same argument, $h_{p_3}^{(3)} \neq \tilde{h}_{p_3}^{(3)}$(since in (a) it pairs $P^{\downarrow}$with $P^{\uparrow}$whereas in (b) it pairs with $P^{\downarrow}$).

Thus, at depth $T = 3$ the **patient embeddings differ across graphs (a) and (b)**. ■

*Consequences for outputs*

- **Node-level:** With any shared prediction head $o_v = \ell\left(h_v^{(T)}\right)$, at least one patient output differs because the inputs to $\ell$differ.
- **Graph-level:** For any injective readout (e.g., sum aggregator) followed by an injective linear map/MLP), the graph-level outputs differ, since the multisets of patient embeddings differ.

Combining Theorem 1 and Theorem 2, KG augmentation can strictly enlarge the hypothesis class and improve identifiability under missing modalities (under the stated assumptions).

#### Supplementary Note 2. Cross-cohort validation, modality ablation and graph-sensitivity analyses

##### Cross-cohort validation

We further evaluated whether the learned progression representation could transfer between independently collected labeled cohorts. In a stricter cross-cohort experiment, models were trained exclusively on PPMI and evaluated on the full external PDBP cohort. This setting is challenging because PPMI and PDBP differ in cohort composition and assessment coverage, and PDBP lacks several non-motor instruments available in PPMI, including SCOPA-AUT, GDS, QUIP, STAI, JOLO, HVLT, LNS, semantic verbal fluency and Symbol–Digit Matching (Supplementary Table 5).

Under this external validation setting, conventional baselines showed limited transferability, with AUROC values close to chance for XGBoost (0.498), LightGBM (0.464) and the plain GNN (0.523). By contrast, KG augmentation improved discrimination (GNN+KG AUROC = 0.743; AUPRC = 0.513), and the full MedStitcher model achieved the best overall external performance on PDBP (AUROC = 0.759; AUPRC = 0.562; Supplementary Table 2). These results indicate that MedStitcher retains predictive signal under cross-cohort distribution shift and that its gains are not limited to within-cohort resampling.

##### Modality ablation analysis

To define which information sources contributed to this performance, we performed modality ablation analyses within the full MedStitcher framework. Removing clinical features caused the largest degradation, reducing pooled AUROC from 0.819 ± 0.040 to 0.661 ± 0.046, indicating that structured clinical phenotyping provides the principal scaffold for rapid progressor discrimination. Removing genetic features also substantially impaired performance, reducing pooled AUROC to 0.712 ± 0.146, whereas removal of MRI features produced a more moderate decline (AUROC = 0.790 ± 0.037). Demographic ablation had the smallest effect on discrimination (AUROC = 0.812 ± 0.047). Together, these results suggest that MedStitcher integrates complementary multimodal information, with clinical assessments acting as the dominant driver and genetic and imaging features providing additional robustness (Extended Data Fig. 2, Supplementary Table 3).

**Graph-construction sensitivity analysis**

Sensitivity analyses further showed that performance was not driven by a narrow graph-construction choice. Removing patient–patient similarity edges entirely degraded pooled performance from AUROC 0.819 ± 0.040 to 0.716 ± 0.114, supporting the importance of patient-level relational structure. However, performance remained stable across moderate graph sparsity settings: top-K values of 10, 20 and 80 yielded pooled AUROC values of 0.810 ± 0.042, 0.796 ± 0.034 and 0.805 ± 0.021, respectively.

Similarly, varying KG message-passing depth from the default two-hop configuration to one or three hops produced only moderate changes in pooled AUROC, with values of 0.784 ± 0.037 and 0.798 ± 0.042, respectively. The default two-hop model provided the best overall balance of discrimination and cohort-level robustness (Extended Data Fig. 2, Supplementary Table 4).

**Supplementary Note 3. Explanatory knowledge graph structure after transfer to Real World Data**

To determine whether the cohort-trained progression signal retained mechanistic structure after transfer to unlabeled real-world EHR data, we applied GNNExplainer to AoU predictions. The resulting explanatory KG was organized around a shared intermediate core rather than dataset-specific peripheral branches, with prominent hubs including HERPUD1, EIF4EBP1, TMED10, CLTB, RAN, MELK and PLEKHM1, and high-weight edges including HLA-DRB1–HLA-DRA, HERPUD1–HLA-DRB1, CTSB–APOE, SREBF1–PLEKHM1 and Generalized anxiety disorder–Anxiety disorder (Extended Data Fig. 3a,b). These structures resolved into three interpretable axes: immune/HLA signaling, lysosomal–proteostatic biology linked to cognitive vulnerability, and neuropsychiatric burden. The HLA-centered module is consistent with genetic evidence implicating HLA class II variation in PD risk[6], whereas the APOE-containing lysosomal/proteostatic neighborhood is consistent with longitudinal evidence linking APOE and lysosomal genetic vulnerability to cognitive decline and dementia in PD[7]. Together, these findings suggest that MedStitcher transfers a biologically structured, hypothesis-generating progression representation to real-world EHR data, rather than relying on arbitrary cohort-specific correlations.

### Supplementary Figure

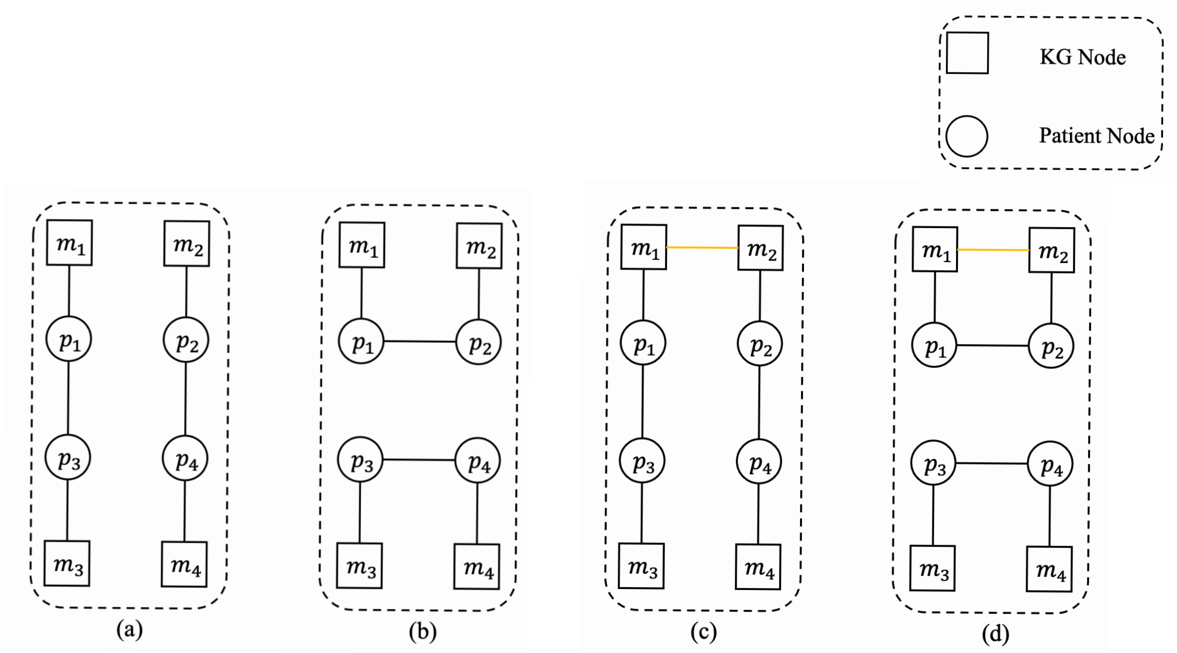


**Supplementary Figure 1**. **Illustration of the theoretical rationale for knowledge-graph (KG) augmentation. a–d**, Toy example illustrating the neighborhood structures used in Theorem 2. Squares denote KG nodes and circles denote patient nodes. Dashed boxes indicate the subgraph extracted for representation learning. **a**, Local patient-centered subgraph $G_S(u)$ for patient $u$, constructed from patient–patient similarity edges and patient–KG connections within the original stitched graph. **b**, Corresponding subgraph $G_S(v)$ for another patient $v$, illustrating that under sparse or partially observed modalities, patient-level neighborhoods may remain difficult to distinguish. **c**, Augmented subgraph $\tilde{G}_{S,KG}(u)$ after KG-based expansion, where shared KG entities (e.g., molecular or phenotype nodes) introduce additional relational paths between patients through structured biomedical context. **d**, Augmented subgraph $\tilde{G}_{S,KG}(v)$, demonstrating how KG-mediated connectivity increases the effective neighborhood overlap and enhances distinguishability between patients with divergent progression trajectories, even under missing or misaligned observed features. This schematic illustrates the mechanism formalized in Theorem 2: KG augmentation enlarges the expressive neighborhood of patient nodes and introduces structured surrogate connections that reduce representational degeneracy under modality incompleteness, thereby improving subtype separability and robustness to cross-cohort distribution shift.

### Supplementary Tables

**Supplementary Table 1. Comparative performance of MedStitcher and baseline models for predicting rapid Parkinson's disease progression across research cohorts.** Red text indicates the best-performing model for each metric within the corresponding evaluation setting.

| **Model** | **Total** | | | | **PPMI** | | | | **PDBP** | | | |
|---|---|---|---|---|---|---|---|---|---|---|---|---|
| Model | AUC | F1 | Precision | Recall | AUC | F1 | Precision | Recall | AUC | F1 | Precision | Recall |
| XGBoost | 0.734 ± 0.027 | 0.365 ± 0.096 | 0.430 ± 0.076 | 0.323 ± 0.118 | 0.680 ± 0.063 | 0.196 ± 0.186 | 0.238 ± 0.227 | 0.187 ± 0.196 | 0.764 ± 0.067 | 0.494 ± 0.137 | 0.554 ± 0.188 | 0.467 ± 0.143 |
| LightGBM | 0.704 ± 0.050 | 0.370 ± 0.097 | **0.433 ± 0.117** | 0.353 ± 0.134 | 0.636 ± 0.089 | 0.226 ± 0.151 | **0.411 ± 0.376** | 0.225 ± 0.196 | 0.746 ± 0.075 | 0.502 ± 0.110 | 0.531 ± 0.120 | 0.489 ± 0.125 |
| GNN | 0.587 ± 0.078 | 0.350 ± 0.036 | 0.255 ± 0.023 | 0.569 ± 0.131 | 0.474 ± 0.170 | 0.120 ± 0.165 | 0.095 ± 0.132 | 0.173 ± 0.241 | 0.382 ± 0.044 | 0.435 ± 0.015 | 0.278 ± 0.012 | **1.000 ± 0.000** |
| GNN+AoU | 0.578 ± 0.117 | 0.290 ± 0.160 | 0.233 ± 0.126 | 0.395 ± 0.219 | 0.587 ± 0.137 | 0.266 ± 0.179 | 0.204 ± 0.144 | 0.404 ± 0.273 | 0.530 ± 0.098 | 0.320 ± 0.141 | 0.292 ± 0.080 | 0.389 ± 0.219 |
| GNN+KG | 0.688 ± 0.039 | 0.343 ± 0.056 | 0.291 ± 0.212 | 0.806 ± 0.301 | 0.623 ± 0.071 | 0.198 ± 0.112 | 0.121 ± 0.073 | 0.691 ± 0.453 | 0.740 ± 0.093 | 0.487 ± 0.101 | 0.360 ± 0.171 | 0.933 ± 0.149 |
| GNN+KG+AoU | 0.773 ± 0.072 | 0.406 ± 0.067 | 0.283 ± 0.083 | **0.824 ± 0.158** | 0.711 ± 0.059 | 0.286 ± 0.031 | 0.191 ± 0.038 | **0.696 ± 0.258** | 0.798 ± 0.170 | 0.578 ± 0.110 | 0.426 ± 0.132 | 0.960 ± 0.055 |
| MedStitcher | **0.819 ± 0.040** | **0.504 ± 0.052** | 0.406 ± 0.039 | 0.673 ± 0.111 | **0.786 ± 0.041** | **0.422 ± 0.098** | 0.322 ± 0.090 | 0.618 ± 0.114 | **0.841 ± 0.129** | **0.629 ± 0.082** | **0.580 ± 0.138** | 0.731 ± 0.163 |

**Supplementary Table 2. Cross-cohort generalization performance for early Parkinson's disease progression prediction (PPMI → PDBP).** Red text indicates the best-performing model for each metric within the corresponding evaluation setting.

| **Model** | **AUC-ROC** | **AUC-PRC** | **F1** | **Precision** | **Recall** |
|---|---|---|---|---|---|

| | | | | | |
|---|---|---|---|---|---|
| XGBoost | 0.498 | 0.299 | 0.177 | 0.233 | 0.143 |
| LightGBM | 0.464 | 0.298 | 0.279 | 0.324 | 0.245 |
| GNN | 0.523 | 0.283 | 0.119 | 0.222 | 0.082 |
| GNN+KG | 0.743 | 0.513 | 0.109 | 0.500 | 0.061 |
| MedStitcher | **0.759** | **0.562** | **0.556** | **0.610** | **0.510** |

**Supplementary Table 3**. **Modality ablation analysis of MedStitcher across research cohorts.** Red text indicates the best-performing model for each metric within the corresponding evaluation setting.

| Modality | Total | | | | PPMI | | | | PDBP | | | |
|---|---|---|---|---|---|---|---|---|---|---|---|---|
| Modality | AUC | F1 | Precision | Recall | AUC | F1 | Precision | Recall | AUC | F1 | Precision | Recall |
| MedStitcher | **0.819 ± 0.040** | **0.504 ± 0.052** | 0.406 ± 0.039 | **0.673 ± 0.111** | **0.786 ± 0.041** | **0.422 ± 0.098** | **0.322 ± 0.090** | **0.618 ± 0.114** | **0.841 ± 0.129** | **0.629 ± 0.082** | 0.580 ± 0.138 | **0.731 ± 0.163** |
| w/o MRI | 0.790 ± 0.037 | 0.490 ± 0.064 | 0.439 ± 0.099 | 0.612 ± 0.206 | 0.750 ± 0.046 | 0.428 ± 0.063 | 0.358 ± 0.089 | 0.585 ± 0.173 | 0.820 ± 0.097 | 0.577 ± 0.107 | 0.578 ± 0.164 | 0.640 ± 0.251 |
| w/o Clinical | 0.661 ± 0.046 | 0.000 ± 0.000 | 0.000 ± 0.000 | 0.000 ± 0.000 | 0.656 ± 0.075 | 0.000 ± 0.000 | 0.000 ± 0.000 | 0.000 ± 0.000 | 0.592 ± 0.081 | 0.000 ± 0.000 | 0.000 ± 0.000 | 0.000 ± 0.000 |
| w/o Genetic | 0.712 ± 0.146 | 0.248 ± 0.175 | 0.227 ± 0.157 | 0.394 ± 0.397 | 0.714 ± 0.074 | 0.205 ± 0.151 | 0.165 ± 0.116 | 0.404 ± 0.407 | 0.721 ± 0.271 | 0.327 ± 0.230 | 0.402 ± 0.305 | 0.384 ± 0.387 |
| w/o Demographic | 0.812 ± 0.047 | 0.462 ± 0.113 | **0.426 ± 0.061** | 0.531 ± 0.203 | 0.777 ± 0.044 | 0.337 ± 0.137 | 0.295 ± 0.105 | 0.411 ± 0.200 | 0.834 ± 0.105 | 0.615 ± 0.133 | **0.615 ± 0.139** | 0.660 ± 0.241 |

**Supplementary Table 4. Sensitivity analysis of patient–patient graph sparsity and message-passing depth.** Red text indicates the best-performing model for each metric within the corresponding evaluation setting.

| Patient-patient graph sparsity | KG size | Total | | | | PPMI | | | | PDBP | | | |
|---|---|---|---|---|---|---|---|---|---|---|---|---|---|
| Top-k edge | hop | AUC | F1 | Precision | Recall | AUC | F1 | Precision | Recall | AUC | F1 | Precision | Recall |
| **40 (default)** | **2 (default)** | **0.819 ± 0.040** | 0.504 ± 0.052 | 0.406 ± 0.039 | 0.673 ± 0.111 | **0.786 ± 0.041** | 0.422 ± 0.098 | 0.322 ± 0.090 | **0.618 ± 0.114** | **0.841 ± 0.129** | 0.629 ± 0.082 | 0.580 ± 0.138 | 0.731 ± 0.163 |
| 0 | 2 | 0.716 ± 0.114 | 0.302 ± 0.167 | 0.297 ± 0.121 | 0.322 ± 0.206 | 0.681 ± 0.161 | 0.200 ± 0.180 | 0.200 ± 0.162 | 0.205 ± 0.206 | 0.692 ± 0.177 | 0.388 ± 0.175 | 0.399 ± 0.096 | 0.444 ± 0.301 |
| 10 | 2 | 0.810 ± 0.042 | **0.562 ± 0.045** | **0.486 ± 0.020** | **0.676 ± 0.126** | 0.781 ± 0.069 | **0.445 ± 0.055** | **0.375 ± 0.035** | 0.564 ± 0.146 | **0.841 ± 0.063** | **0.697 ± 0.089** | **0.626 ± 0.023** | **0.800 ± 0.187** |
| 20 | 2 | 0.796 ± 0.034 | 0.466 ± 0.070 | 0.449 ± 0.087 | 0.510 ± 0.141 | 0.757 ± 0.034 | 0.344 ± 0.090 | 0.315 ± 0.090 | 0.395 ± 0.120 | 0.830 ± 0.106 | 0.615 ± 0.228 | 0.646 ± 0.291 | 0.633 ± 0.259 |
| 80 | 2 | 0.805 ± 0.021 | 0.477 ± 0.127 | 0.460 ± 0.070 | 0.523 ± 0.193 | 0.762 ± 0.047 | 0.341 ± 0.089 | 0.349 ± 0.085 | 0.360 ± 0.126 | 0.828 ± 0.119 | 0.622 ± 0.224 | 0.581 ± 0.173 | 0.700 ± 0.308 |
| 40 | 1 | 0.784 ± 0.037 | 0.446 ± 0.098 | 0.427 ± 0.089 | 0.492 ± 0.149 | 0.741 ± 0.027 | 0.309 ± 0.114 | 0.273 ± 0.083 | 0.398 ± 0.180 | 0.829 ± 0.086 | 0.641 ± 0.140 | 0.711 ± 0.209 | 0.593 ± 0.116 |
| 40 | 3 | 0.798 ± 0.042 | 0.473 ± 0.090 | 0.454 ± 0.150 | 0.521 ± 0.084 | 0.760 ± 0.061 | 0.401 ± 0.107 | 0.369 ± 0.153 | 0.475 ± 0.107 | 0.833 ± 0.092 | 0.572 ± 0.139 | 0.605 ± 0.201 | 0.569 ± 0.141 |

**Supplementary Table 5. Clinical variables used for PD subtyping**

| Category | Data | Description | PPMI | PDBP |
|---|---|---|---|---|
| Motor assessment | MDS-UPDRS Part II[8] | Self-administered questionnaire of motor experiences of daily living. We used all items. | X | X |
| | MDS-UPDRS Part III[8] | Motor examination provided by rater. We used all items with medication "OFF". | X | X |
| | Schwab-England activities of daily living score | Measure of the abilities of individuals living with PD relative to a completely independent situation. | X | X |
| Non-motor assessment | MDS-UPDRS Part I[8] | Non-motor experiences of daily living. We used all items. | X | X |
| | Scales for Outcomes in Parkinson's disease-Autonomic (SCOPA-AUT) [9] | The SCOPA-AUT was developed to evaluate autonomic symptoms. We used scores of the 7 domains, including gastrointestinal, urinary, cardiovascular, thermoregulatory, pupillomotor, and sexual. | X | |
| | Geriatric depression scale (GDS) [10] | Measure of depression in older adults. | X | |
| | Questionnaire for Impulsive-Compulsive Disorders in Parkinson's disease (QUIP)[11] | Measure of severity of symptoms and support a diagnosis of impulse control disorders and related disorders in PD. We used all items. | X | |
| | State-Trait Anxiety Inventory (STAI)[12] | The measure of trait and state anxiety. We used the STAI-Trait and STAI-State sub-scores. | X | |
| | Benton Judgment of Line Orientation (JOLO)[13] | A standardized measure of visuospatial judgment. We used the crude score and MOANS normative scores. | X | |
| | Hopkins Verbal Learning Test (HVLT)[14] | A memory test with six equivalent forms. | X | |
| | Letter-number sequencing (LNS) | A subset of Wechsler adult intelligence scale, measuring working memory, attention, mental control | X | |
| | Montreal Cognitive Assessment (MoCA)[15] | A screening assessment for detecting cognitive impairment. We used the visuospatial, naming, attention, language, delayed recall, abstraction, and verbal fluency sub-scores, and total MoCA score. | X | X |
| | Semantic verbal-language fluency test[16] | Assessment of semantic knowledge, retrieval ability, and executive functioning. We used the sub-scores in terms of animals, vegetables, and fruits. | X | |
| | Symbol–Digit Matching (SDM)[17] | A neuropsychological test that examines a person's attention and speed of processing. | X | |
| | Epworth Sleepiness Score (ESS)[18] | Measure of daytime sleepiness. We used all items. | X | X |
| | REM sleep behaviour disorder (RBD)[19] | A questionnaire for RBD. We used all items. | X | X |
| | Cranial Nerve Examination | A kind of neurological examination that is used to identify problems with the cranial nerves. We used the 9 components. | X | |
| PD medication | Levodopa equivalent daily dose | Levodopa equivalent daily dose | X | |

Abbreviations: MDS-UPDRS = Movement Disorders Society–revised Unified Parkinson's Disease Rating Scale.
